# HPV Self-Sampling in Cervical Screening: A Rapid Review

**DOI:** 10.64898/2026.06.12.26354841

**Authors:** Novatus Apolinary Tesha, Clareece Nevill, Martin Taylor-Rowan, Ryan Mulholland, Terry Quinn, Anna Noel-Storr, Alex J Sutton, Olivia Wu

**Author notes:** **Corresponding Author Novatus Apolinary Tesha:** /, **Address:** Health Economics and Health Technology Assessment (HEHTA) School of Health & Wellbeing, College of Medical, Veterinary & Life Sciences University of Glasgow, Clarice Pears Building, 90 Byres Road, Glasgow G12 8TB.

## Abstract

**Introduction:** Cervical cancer is the fourth largest cause of cancer deaths in women. HPV self-sampling could increase uptake of cervical screening. This rapid review aimed to determine the accuracy, concordance, uptake and acceptability of self-sampling over clinician-collected samples in high income countries.

**Method:** We followed Cochrane Rapid Reviews Methods. Top-up of 4 systematic reviews and meta-analyses was performed. Narrative data synthesis was conducted and meta-analysis where applicable. Databases searched were MEDLINE, EMBASE, CENTRAL and clinical trial registries. Risk of bias was assessed using AMSTAR 2, QUADAS, the Cochrane Risk of Bias (RoB), or the Nudelman and Otto, 2020 tool, depending on the study type.

**Findings:** The review included 39 studies for accuracy, 38 studies for concordance, 37 uptake and 48 studies for acceptability. Self-sampling has similar accuracy as clinician-collected samples when PCR-based assays are used. The overall agreement of self-sampling and clinician-collected samples was 87.1%(95%CI;85.6-88.6) with a kappa value of 0.70(95%CI;0.67-0.73). Mail-to-all strategies had higher uptake with participation differences of 11.3%(95%CI:8.4-14.2) in the intention-to-treat analysis and 7.7%(95%CI:4.7-10.8) in the per protocol analysis. Self-sampling is acceptable to non-attendees (91%(95%CI;85.3-94.6).

**Conclusion and Recommendation:** Self-sampling shows good performance on the four clinical effectiveness indicators of accuracy, concordance, uptake and acceptability.

## Introduction

Cervical cancer is the fourth commonly diagnosed cancer among women in the world (1). More than 95% of cervical cancer is caused by persistent infection with Human Papillomavirus (HPV)(2). There are more than 100 HPV genotypes, categorized into high-risk (hrHPV) and low risk/non-oncogenic strains. There at least 14 types that can cause cancer some of which are hrHPV16, 18, 31 and 33(3). hrHPV 16 and 18 together cause about 70% of cervical cancer(3). Cervical cancer takes a long time to develop. It passes through a precancerous stage which is termed as cervical intraepithelial neoplasia (CIN). CIN is categorized according to the severity of dysplasia as CIN1 (low grade) and CIN2+ (high grade) (4). Most of women (90%) who get infection with HPV, the virus clear themselves within 24 months, however, for a few the virus persist and can lead into cervical cancer(2). It takes between 10 to 20 years for women infected with HPV with strong immunity to develop cancer. This long time lag provides an opportunity for early detection of precancerous lesions via regular screening followed by immediate treatment (5).

A range of diagnostic HPV-DNA tests and sampling methods are available, and samples may be self-collected from the vagina, as an alternative to collection from the cervix by a healthcare professional(6). The traditional screening methods has been cytology, the Papanicolaou test (pap smear), this test has been used in high income countries (HICs) and is responsible for the low cervical cancer burden in HICs(5). However, in 2021 WHO published a new guideline which recommends countries to use HPV DNA as a primary screening method in the place of pap smear and VIA(7). HPV DNA can be done by self-sampling or clinician collected.

Most high-income countries have national screening programs; however, the uptake of screening is still suboptimal. As a result, cervical cancer is most frequently diagnosed in those who are either under screened or never screeners (8,9). The non-attendance in regular screening is attributed to time constraints, inconveniences, lack of awareness, anxiety, pain, embarrassment and physical discomfort during specimen collection. Service issues may also present barriers to participation, such as a lack of suitable appointment times, or nearby clinics(10). HPV DNA self sampling may contribute to attracting higher response and overcome challenges associated with clinician collected samples(11) Several countries, including France, Sweden and Australia, have incorporated HPV DNA self-sampling into their national screening programmes, either as a primary screening approach, or as a method targeted at under screened individuals.

Recent global systematic reviews and meta-analysis on the accuracy, concordance, uptake and acceptability of the HPV self-sampling found that HPV self-sampling has similar accuracy and concordance with the clinician collected samples when polymerase chain reaction (PCR) based assay are used and it has a higher uptake(9,12). It was more acceptable than the clinician collected samples(11,13). However, there were heterogeneity (9,12–14). It was recommended that country or regions should conduct pilots assessing feasibility, effectiveness and cost effectiveness before rolling out HPV DNA self-sampling(15). In this regards, different countries, including the UK, piloted HPV self-sampling (You Screen)(16). As the findings of these pilots are coming out, and HICs countries are moving towards HPV DNA self-sampling, we aim to pool the findings from HICs to contextualise the findings of self-sampling to these settings and inform the adoption of this HPV DNA self-sampling in these settings. Therefore, the aim of this rapid review is to compare the clinical performance of HPV self-sampling on four aspects: accuracy, concordance, uptake and acceptability compared to clinician-collected samples.

## Methods

The protocol for this review was registered in PROSPERO with registration CRD42024573464. The methods of this rapid review followed the Cochrane Rapid reviews Method Groups guidance (17–21). It also accounts for the methodological challenges of diagnostic reviews (22). A detailed explanation of the methodological approach to the review is included within the Supplementary Materials.

### Study Design

This is the rapid review on the four clinical aspects of HPD DNA self-sampling compared clinician collected samples. It answers the following four clinical questions on accuracy, concordance, uptake and acceptability of the HPV DNA self-sampling compared to clinician collected samples.

1. What is the accuracy of HPV testing in self-collected samples compared with health professional collected samples, and does this vary according to eligible women and test characteristics? *(accuracy*)
2. What is the level of concordance between HPV-DNA testing in self-collected samples and clinician/health professional collected samples, and does this vary according to eligible women and test characteristics? (*concordance)*
3. What is the uptake of cervical screening by HPV self-sampling method when compared to health professional sampling method in non-attenders with those offered health professional sampling, and does this vary according to eligible women and test characteristics? *(uptake*)
4. Are HPV self-sampling screening strategies acceptable to those that have not attended the regular cervical screening programme, and does this vary according to eligible and test characteristics? (acceptability)

This rapid review updates the already existing reviews on the research questions with new evidence published up to March 2024. The reference reviews were Arbyn et al (15) for accuracy, Arbyn(15,23) for concordance, Arbyn et al (15) and Costa et al(9) for uptake and Nelson et al(13) for acceptability. These reviews formed the basis for data extraction.

Accuracy and concordance questions focused on women eligible for cervical cancer screening, while the latter two focused on women who under/never screeners. The intervention is the HPV DNA self*-*sampling and the comparators been HPV testing on healthcare professional-collected sample. This rapid review included only studies conducted in High Income Countries

### Literature Search

A comprehensive description of the search strategy is provided within the Supplementary Materials. Briefly, the databases used included Medline, Embase, Central and clinical trial registries. The respective systematic reviews upon which each search strategy is based are reported, with the search strategies detailed in *Supplemental table 1.* The start dates for the searches were intended to allow for a three-month overlap with the end dates of the search strategies reported in the existing reviews, to ensure all relevant studies were identified. Indeed, the start date was 1^st^ January 2018 for the search strategies for accuracy, concordance and uptake questions (overlap with Arbyn et al. 2018(14)) and 1^st^ December 2014 (overlap with Nelson et al. 2015(13)), respectively, the end date for all searches was March 2024. The existing systematic reviews were otherwise assumed to have captured all relevant studies prior to the start dates in the searches conducted for this rapid review.

### Eligibility Criteria

Abstracts, conference proceedings and non-English language studies were excluded from the review. We included studies which had a minimum of 1000 participants for uptake.

### Screening Process

All studies fulfilling the eligibility criteria were included in the review. Screening of abstracts were conducted by two independent reviewers (NT and RM). Full text records were screened by one reviewer and validation of excluded records (20%) was undertaken by a second reviewer. All discrepancies were resolved by consensus and/or a third reviewer.

### Data Extraction

Data extraction from individual reviews and studies were carried out by a single reviewer. Where feasible, data were extracted from existing systematic reviews. Data extraction was then completed for additional studies identified in the searches that have not been captured in prior reviews.

### Statistical Analysis

The analysis for all questions followed the Cochrane Handbook for Systematic Review of Diagnostic Test Accuracy where appropriate: Meta-analyses were primarily conducted in R(24) using the {meta}(25)or {metafor} package(26). Forest plots were produced to investigate potential heterogeneity in meta-analyses. Where data permitted, meta-regressions were conducted. (*See appendix)*

### Quality of Included Studies

The quality of the reference reviews was assessed using AMSTAR 2. The accuracy studies were assessed using the QUADAS tool for both studies included in the review and post review studies. The Cochrane Risk of Bias(RoB) was used by the uptake reference review and post review studies. The risk of bias of the studies in the acceptability study were assessed using Nudelman and Otto, 2020 tool Risk of Bias Utilized for Surveys Tool (ROBUST)(27). The RoB for concordance studies were not assessed since there is no a validated tool for assessing concordance studies.

## Results

There were 1,319 studies identified from searching databases and registries, of which 88 were eligible (*Figure 1*). Combined with 92 studies identified from systematic reviews, this gave a total of 180 studies across the four clinical questions.

**Figure 1:**
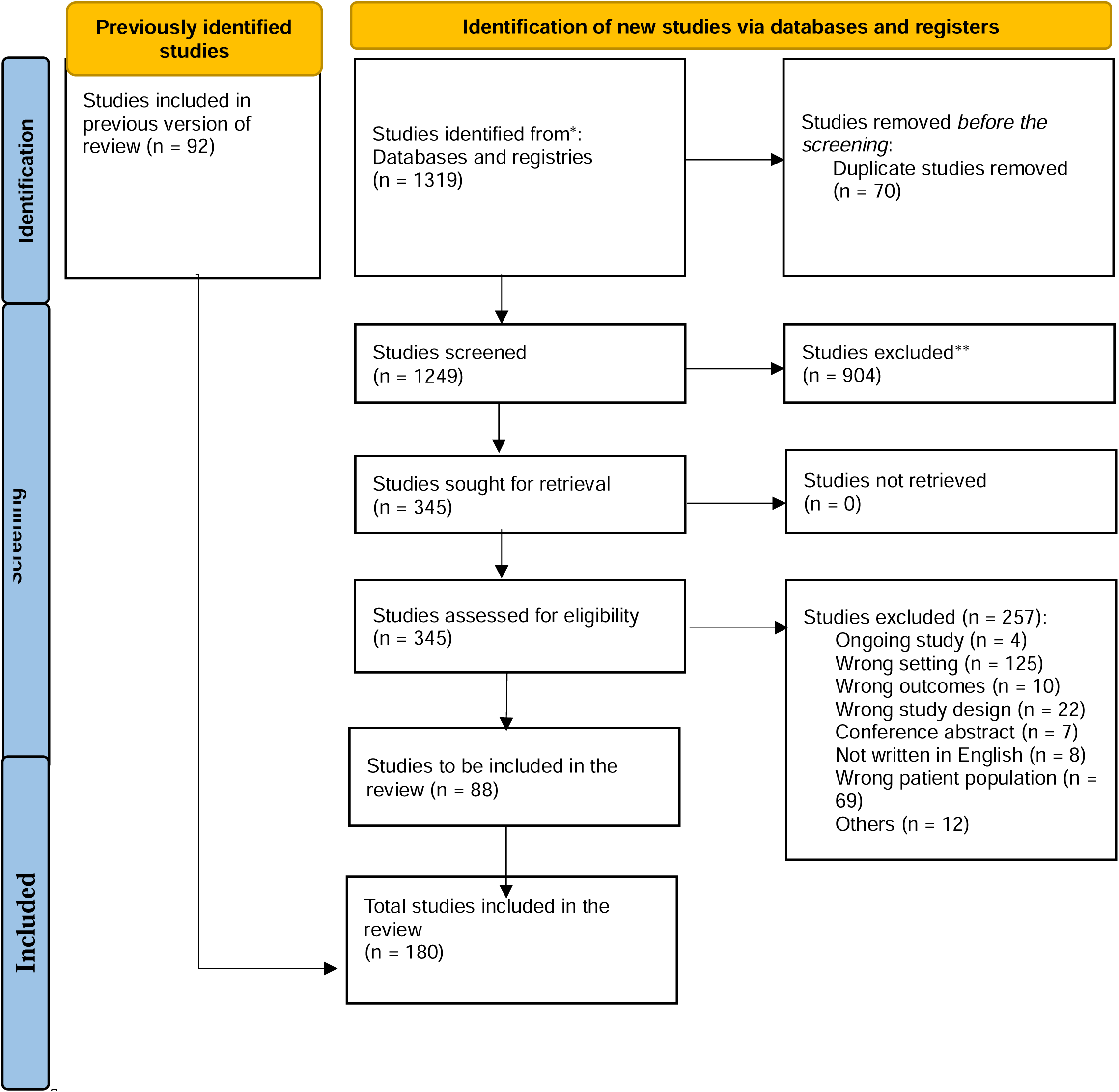
PRISMA Flow Chart for the Included Studies. *The included studies were 39 studies for accuracy, 38 studies for concordance, 37 uptake and 48 studies for acceptability. The studies were not mutually exclusive.*

### Characteristics of Included Studies

#### Accuracy of HPV testing in self-collected samples compared with health professional-collected samples

The accuracy question review included 39 studies – 18 studies from the referenced review Arbyn et al 2018 (15) and 21 from the top-up search (*Supplementary Table 2*). The studies were conducted in 13 different high-income countries. 7 studies included women who were attending primary screening, 30 studies included women who were referred for colposcopy and 1 study included patients with cervical premalignant lesion/carcinoma diagnosis or carcinoma suspicion(28) and another study included outpatients with abnormal cytology and requiring colposcopy and biopsy/negative for intraepithelial lesions or malignancy NILM/HPV-positive patients(29). The more common self-sampling devices reported in these studies was brush (15). The assay used in these studies included PCR (28).

#### The level of concordance between HPV-DNA testing in self-collected samples and health professional collected samples in cervical screening non-attenders

The concordance question included 50 studies – 26 from reference review (12 studies had no outcome and hence not included in this study-*Supplemental table 3*) and 24 from the top-up search. The studies were conducted in 15 different countries, and the number of participants in the studies ranged from 25 to 4,617. Most of the studies included women who were referred for colposcopy clinic (26). The self-sampling device used included brush (14) (*Supplemental table 4*).

#### Uptake of cervical screening by HPV self-sampling method when compared to health professional sampling method in non-attenders with those offered health professional sampling

The uptake question included 38 studies – 26 articles from the existing review (One had no outcome - *Supplemental Table 5*) and 12 studies from the top-up articles. These studies were from 17 high-income countries. The number of participants ranged from 529 to 57,717 in the self-sampling arm and 261 to 23,632 in the control arm. Almost all studies used either opt-in (7), mail-to-all all (20), or a combination of these two self-sampling strategies (10). The sampling devices used were brush (11), swab (13) and lavage (4). Most of the studies reported both per protocol (PP) and intention to treat (ITT) analyses (30) (*Supplemental Table 6*).

#### Acceptability of HPV self-sampling screening strategies to those that have not attended the regular cervical screening programme

The acceptability question had 53 articles: 22 from the review (5 studies had no outcome *Supplemental Table 7*) and 31 from the post-review top-up search. The studies were from 19 different countries. The number of participants ranged from 31 – 9,484. The self-sampling devices included in the studies are brush (11), swab (17), lavage (3) and tampon (1) (*Supplemental Table 8*).

### Risk of Bias/ Quality Assessment of the Included Studies

Most of the studies had low risks on items assessed on the accuracy except on the patient selection-applicability concerns only one study that had a low risk of bias with the rest of studies having high risk *(supplemental Table 9).* All the assessed studies on uptake had a serious/high risk of bias. *(Supplemental Table 10).* For the acceptability question, out of 48 studies accessed for quality, 46 studies scored 4 points or below. Most of the studies did not report on the items in the assessment tool *(Supplemental Table 11*).

### Synthesis of Results

*Accuracy of HPV testing in self-collected samples compared with health professional-collected samples* Of the 39 studies, only 13 had data in a format needed for meta-analysis (*Supplementary Table 2)*.

Table 1 gives the pooled estimates for absolute sensitivity and specificity measures for using self-sampling and healthcare professional sampling across different screening groups. Figure 2 gives a forest plot of the absolute difference in accuracy for self-sampling versus healthcare professional sampling across the screening groups. For the largest group, colposcopy referral for CIN2+, the absolute difference was −5.5% (95% CI: −16.2% to 5.2%) and 4.5% (95% CI: −12.7% to 21.7%) for sensitivity and specificity respectively.

**Figure 2:**
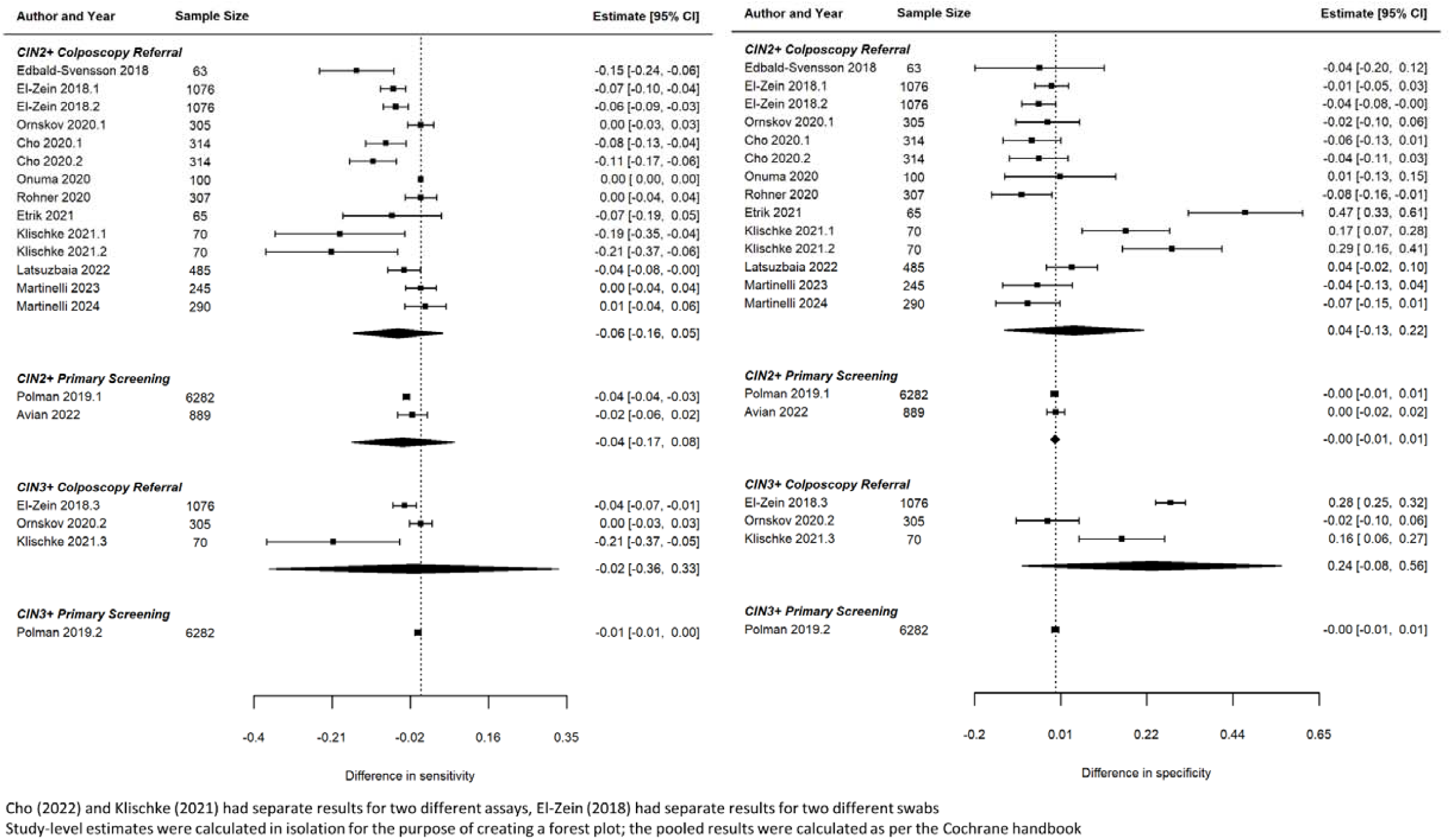
Forest plot of absolute difference in sensitivity and specificity of self-sampling versus healthcare professional sampling for hrHPV screening. *The level of concordance between HPV-DNA testing in self-collected samples and health professional collected samples in cervical screening non-attenders.*

**Table 1:**
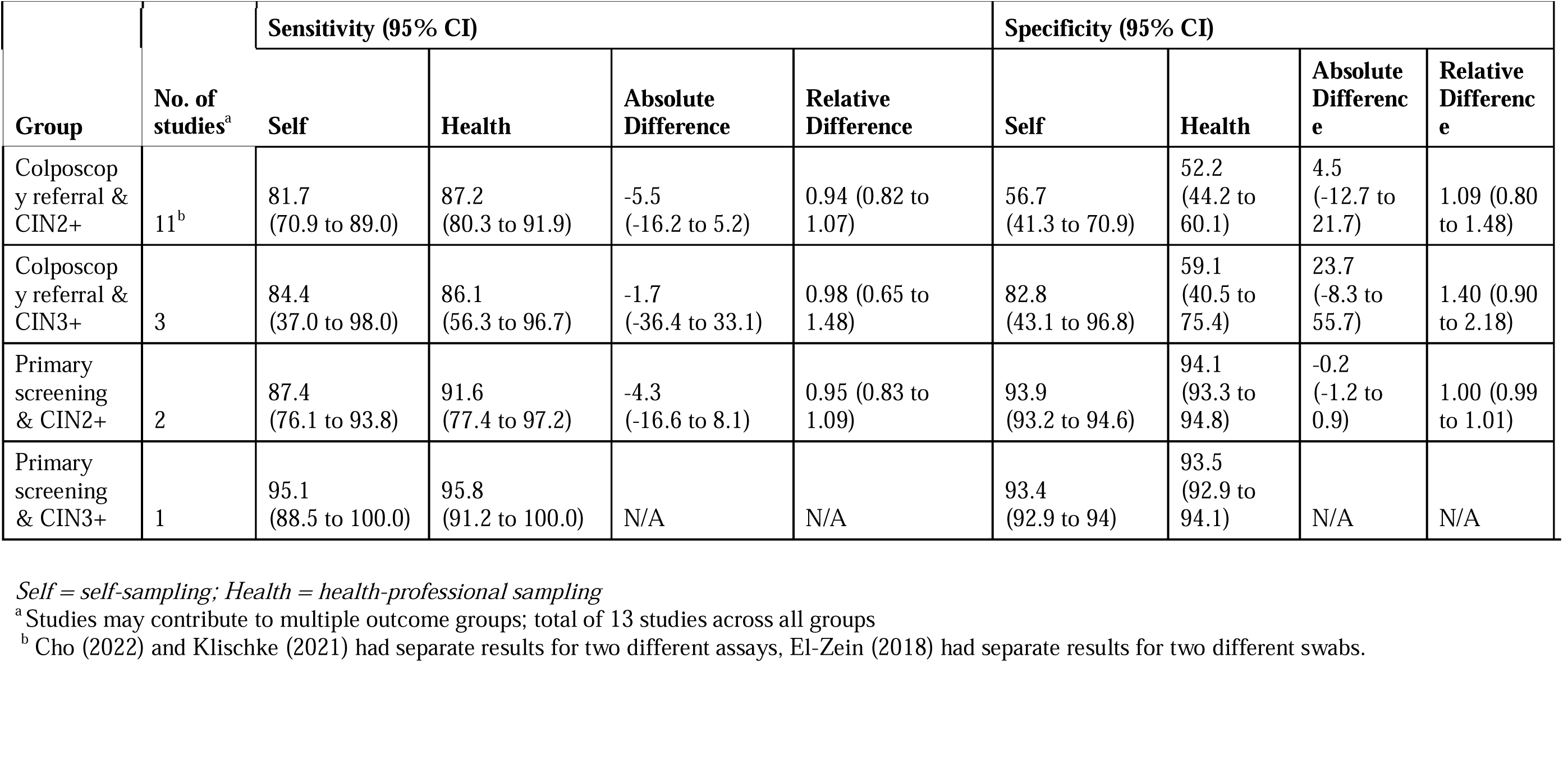
Pooled Estimates for Absolute Accuracy Measures.

| Group | No. of studies<sup>a</sup> | Sensitivity (95% CI) |  |  |  | Specificity (95% CI) |  |  |  |
| --- | --- | --- | --- | --- | --- | --- | --- | --- | --- |
|  |  | Self | Health | Absolute Difference | Relative Difference | Self | Health | Absolute Difference | Relative Difference |
| Colposcopy referral & CIN2+ | 11<sup>b</sup> | 81.7<br>(70.9 to 89.0) | 87.2<br>(80.3 to 91.9) | -5.5<br>(-16.2 to 5.2) | 0.94 (0.82 to 1.07) | 56.7<br>(41.3 to 70.9) | 52.2<br>(44.2 to 60.1) | 4.5<br>(-12.7 to 21.7) | 1.09 (0.80 to 1.48) |
| Colposcopy referral & CIN3+ | 3 | 84.4<br>(37.0 to 98.0) | 86.1<br>(56.3 to 96.7) | -1.7<br>(-36.4 to 33.1) | 0.98 (0.65 to 1.48) | 82.8<br>(43.1 to 96.8) | 59.1<br>(40.5 to 75.4) | 23.7<br>(-8.3 to 55.7) | 1.40 (0.90 to 2.18) |
| Primary screening & CIN2+ | 2 | 87.4<br>(76.1 to 93.8) | 91.6<br>(77.4 to 97.2) | -4.3<br>(-16.6 to 8.1) | 0.95 (0.83 to 1.09) | 93.9<br>(93.2 to 94.6) | 94.1<br>(93.3 to 94.8) | -0.2<br>(-1.2 to 0.9) | 1.00 (0.99 to 1.01) |
| Primary screening & CIN3+ | 1 | 95.1<br>(88.5 to 100.0) | 95.8<br>(91.2 to 100.0) | N/A | N/A | 93.4<br>(92.9 to 94) | 93.5<br>(92.9 to 94.1) | N/A | N/A |

*Self* = self-sampling; *Health* = health-professional sampling
<sup>a</sup> Studies may contribute to multiple outcome groups; total of 13 studies across all groups
<sup>b</sup> Cho (2022) and Klischke (2021) had separate results for two different assays, El-Zein (2018) had separate results for two different swabs.

Self-sampling device and setting did not give a significant effect on the absolute difference for colposcopy referral CIN2+ (LR test p-value = 0.143 and 0.984, respectively). Assay methods were all target-amplification methods regarding colposcopy referral CIN2+. Other groupings were not tested for test characteristic effects due to the small number of studies.

None of the 39 studies identified had data regarding numbers of samples that could not be determined/needed a second sample or data regarding the number of women with a positive test result attending clinics for further investigation.

There were ten studies not included in the meta-analysis due to the lack of information (*Supplemental table 4*). This resulted in 28 studies with at least one concordance outcome that were included in the meta-analysis. The overall agreement was 87.1 (85.6 to 88.6) and the Kappa 0.70 (0.67 to 0.73) –(***Table 2***)

**Table 2:** Pooled Estimates for Concordant Outcomes.

| Outcome | Subgroup** | No. of studies* | All results |
| --- | --- | --- | --- |
| Overall agreement (%) | All | 25 | 87.1 (85.6 to 88.6) |
|  | Clinical setting | 18 | 86.1 (84.0 to 88.0) |
|  | Home setting | 4 | 90.0 (88.0 to 91.6) |
| Kappa | All | 25 | 0.70 (0.67 to 0.73) |
|  | Clinical setting | 18 | 0.73 (0.70 to 0.76) |
|  | Home setting | 4 | 0.62 (0.57 to 0.67) |
| Test positivity rate ratio | All | 12 | 0.97 (0.89 to 1.04) |
|  | Swab & TA assay | 3 | 1.03 (0.99 to 1.06) |
|  | Lavage & TA assay | 1 | 1.24 (1.10 to 1.40) |
|  | Brush & TA assay | 6 | 0.95 (0.90 to 1.00) |
|  | Brush & SA assay | 1 | 0.66 (0.56 to 0.77) |
|  | Brush & RNA assay | 1 | 0.98 (0.90 to 1.06) |
| Positive agreement (%) |  | 17 | 85.5 (81.6 to 88.7) |
| Negative agreement (%) | All | 17 | 82.3 (74.9 to 87.9) |
|  | Clinical setting | 13 | 86.8 (83.6 to 89.5) |
|  | Home setting | 1 | 52.3 (47.1 to 57.5) |
| Positive concordance (%) |  | 13 | 77.0 (70.7 to 82.1) |
| Negative concordance (%) |  | 13 | 74.6 (70.8 to 78.1) |

\* Many studies gave multiple results (e.g. different assays, devices)
\*\* Only reported where the inclusion of the respective variable gave a significant (<0.05) result

The level of overall agreement was found to be higher among home setting than clinical setting; however, this was in direct contrast to that was observed with the kappa measure. The negative agreement and test positivity ratio differed across the self-sampling devices. The negative agreement also differed on the self-sampling settings and test positivity ratio differed across the self-sampling test assay.

*Supplemental Figure 1* and *Figure 2* show forest plots of overall agreement and kappa respectively. The included studies reported overall agreement ranging from 77% to 96% and kappa value ranging from 0.47 to 0.86. There was substantial heterogeneity amongst the studies.

Regarding overall agreement, test assay gave no significant effect (p = 0.292), while self-sampling device gave a borderline significant effect (p = 0.046). However, the only device that gave a significant result was ‘tampon’ which was only informed by one study. There was a statistically significant effect regarding clinical setting (p = 0.008) where overall agreement was higher for tests taken in a home setting (90% agreement (95% CI: 88% to 92%) versus 86% agreement (95% CI: 84% to 88%) (*Supplemental Figure 3*). Regarding kappa, self-sampling device and test assay gave no significant effect (p = 0.948 and p = 0.139, respectively). There was a statistically significant effect regarding clinical setting (p <0.001) where kappa was higher for tests taken in a clinical setting (0.73 (95% CI: 0.70 to 0.76) versus 0.62 (95% CI: 0.57 to 0.67)) (*Supplemental Figure 4)*, which was in direct contrast to the result found for overall agreement.

Negative agreement was also affected by setting of the test (Clinical setting 86.8% (95% CI: 83.6% to 89.5%) versus home setting 52.3% (95% CI: 47.1% to 57.5%), p <0.001) and the test positivity rate ratio was jointly affected by self-sampling device and assay method (p <0.001, see table 2 for effect sizes) (***Table 2***). Other outcomes were not affected by the other characteristics tested.

#### Uptake of cervical screening by HPV self-sampling method when compared to health professional sampling method in non-attenders with those offered health professional sampling

Eight studies were not included in the meta-analysis (*Supplemental Table 6*) leaving 29 studies. *Table 3* shows the percentage of women having a hrHPV test done with a self-sample, separately for those who received a self-sampling kit mailed to their home (mail-to-all) and those having to request a self-sampling kit (opt-in). Overall, the participation rate is higher amongst self-sampling compared with controls.

**Table 3:** Absolute and Relative participation in self-sampling and/versus control arms.

| Invitation scenario | No. of studies | Absolute participation |  | Participation difference % (95% CI) | Relative participation (95% CI) |
| --- | --- | --- | --- | --- | --- |
|  |  | Self-sampling % (95% CI) | Control % (95% CI) |  |  |
| Per protocol |  |  |  |  |  |
| Mail-to-all | 26* | 17.7 (15.0 to 20.8) | 9.1 (6.9 to 12.0) | 7.7 (4.7 to 10.8) | 1.94 (1.48 to 2.55) |
| Opt-in | 10* | 8.5 (6.3 to 11.4) | 9.8 (6.5 to 14.4) | -2.3 (-6.5 to 2.0) | 0.88 (0.51 to 1.52) |
| Intention-to-treat |  |  |  |  |  |
| Mail-to-all | 28* | 23.0 (20.2 to 26.0) | 10.0 (7.4 to 13.2) | 11.3 (8.4 to 14.2) | 2.34 (1.87 to 2.93) |
| Opt-in | 10* | 15.2 (11.5 to 19.8) | 9.8 (6.5 to 14.4) | 5.0 (1.4 to 8.6) | 1.56 (1.09 to 2.24) |

\*Giorgi-Rossi (2011) & Giorgi-Rossi (2015) had two control groups (one with cytology, and another with HPV testing). Kellen (2018) also had two control groups (with and without recall letters).

The pooled participation was higher in the mail-to-all self-sampling strategies compared to control. This was also observed when comparing opt-in strategy with control in the intention-to-treat analysis; however, no statistically significant difference was observed in the per protocol analysis. Overall, the absolute participation rate was greater in the intention-to-treat analysis than in the per protocol analysis.

Absolute participation is the number of responders divided by the total number of individuals in the respective group. Participation difference is the difference between these absolute percentages (i.e. self-sampling – control), and relative participation is the absolute percentage of responders in the self-sampling group divided by the absolute percentage of responders in the control group.

The difference in participation rates of each study included in meta-analysis is shown in *Supplemental Figure 5*.

For the mail-to-all invitation strategy, the time between the invite and a health professional taking the sample affected the participation difference (difference increased by 1.0% (95% CI: 0.1% to 1.8%) and 1.2% (95% CI: 0.4% to 2.0%) per month under the per protocol and intention-to-treat analysis respectively (both estimated from 20 study data points)). No other tested characteristics gave a significant effect for both per protocol and intention-to-treat analyses.

For the opt-in invitation strategy, the use of reminders and time between invite and a health professional taking the sample increased the participation difference by 7.1% (95% CI: 0.5% to 13.6%) and 1.2% (95% CI: 0.5% to 2.0%) (per month, estimated from 10 study data points) respectively under the intention-to-treat analysis. No other tested characteristics gave a significant effect. None of the tested characteristics gave a significant effect for the opt-in invitation strategy under the per protocol analysis.

When incorporating the above characteristics, the heterogeneity did not significantly improve (i.e. I^2^ remained above 96% for all groups).

The pooled proportion of unsatisfactory samples taken by the self-sampling group, their adherence to follow-up, and the CIN2+ detection per 1000 women invited are show in *Supplemental Table 12*. Due to only two studies reporting such information for control arms, pooled relative rates could not be estimated.

#### Acceptability of HPV self-sampling screening strategies to those that have not attended the regular cervical screening programme

All 48 studies were included across the meta-analyses, but studies rarely had data for all the outcomes presented (e.g. some only presented data regarding reasons for (dis)liking self-sampling). The pooled estimates for the acceptability outcomes are shown in *Table 4*. It found that 91% of women are generally accepting of self-sampling, with 74.4% and 59.5% stating preference for doing it at home and doing it themselves rather than a healthcare setting/professional respectively.

**Table 4:**
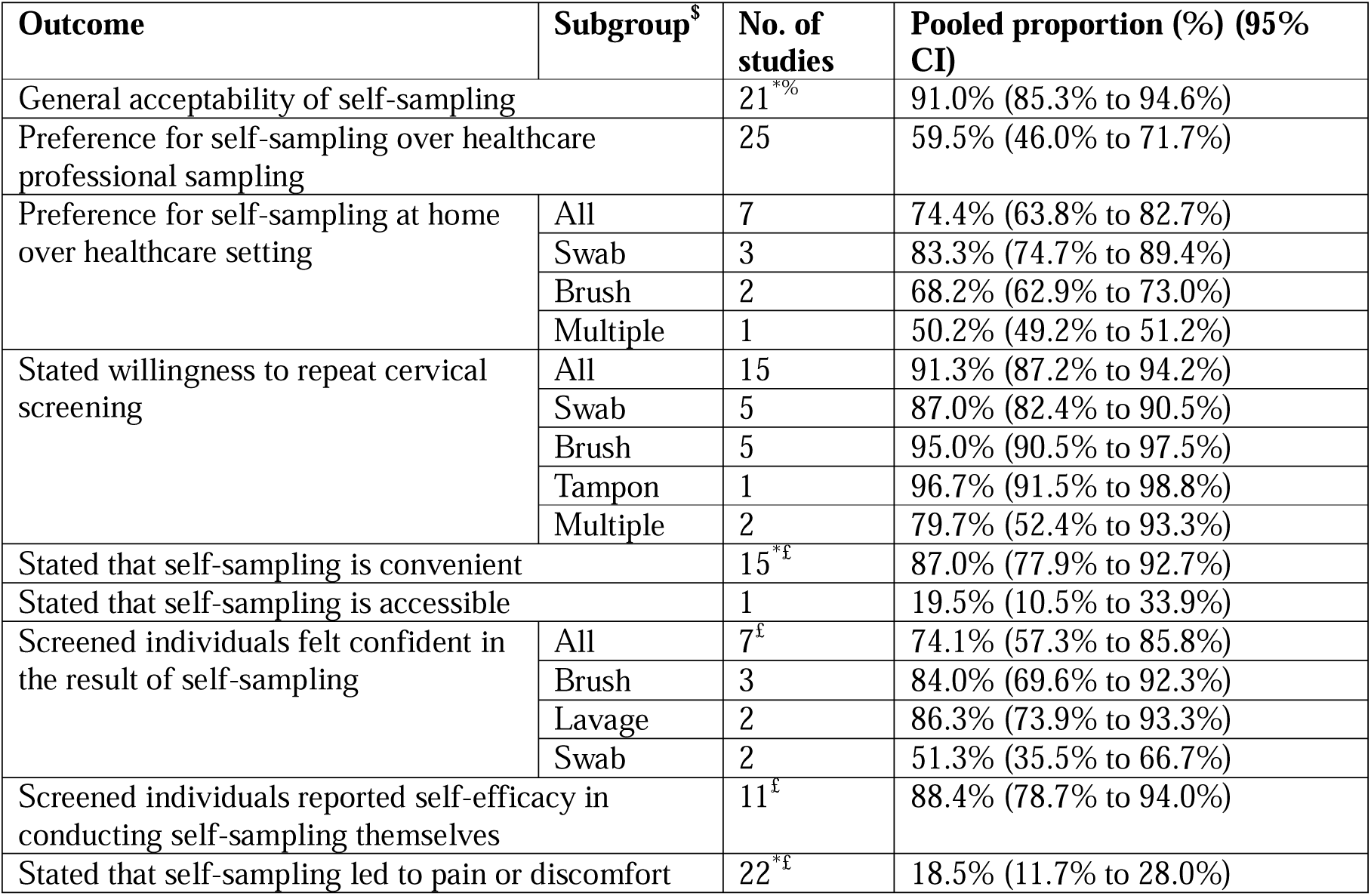
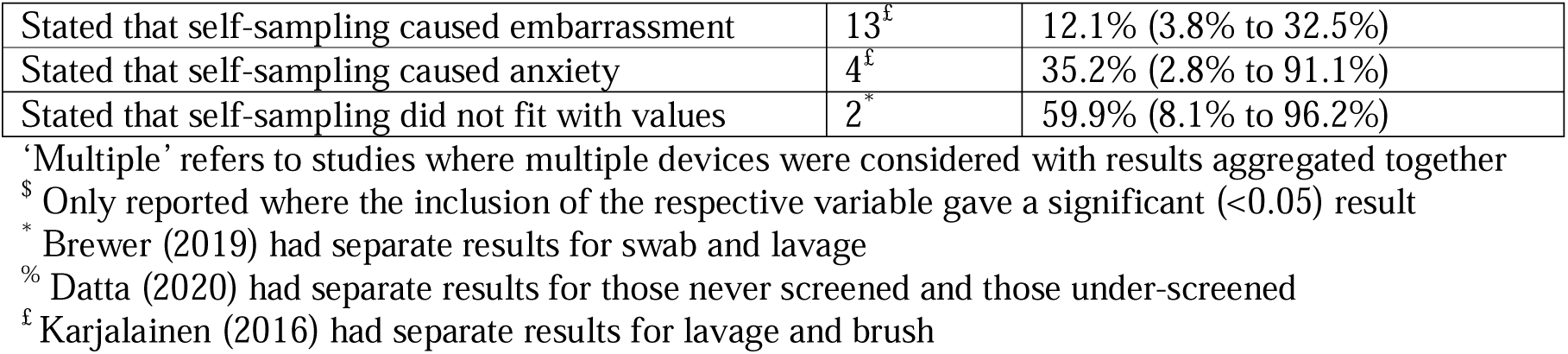
Pooled Analysis for Acceptability Outcomes.

*Supplemental Figure 6* and *Figure 7* illustrate high heterogeneity regarding the general acceptability of self-sampling and its preference over healthcare professionals respectively. *Figure 8* shows consistently high proportions of general acceptability in earlier years, with wide variation in later years. Sampling device was not found to affect general acceptability or preference for self-sampling (p = 0.118 and 0.799, respectively).

High heterogeneity was also observed among the lesser reported acceptability outcomes. Self-sampling device was tested for potential effects, for which only preferences for home setting, willingness to repeat, and individuals feeling confident of the results gave a significant result (*Table 4)*. There were insufficient data in a consistent format for ethnicity or age to be considered in a quantitative manner.

Preference for self-sampling at home over healthcare setting differed across self-sampling device and invitation strategy. *Supplemental Figure 8* and *Figure 9* shows that the preference for a home setting was higher for swabs (83% (95% CI: 75% to 89%) versus 68% (95% CI: 63% to 73%) for brush and 50% (95% CI: 49% to 51%) for combinations, p < 0.001) and higher when offered in a healthcare setting respectively (72% (95% CI: 61% to 81%) versus mail-to-all preference of 50% (95% CI: 49% to 51%), p=0.020). It was not possible to analyse device and invitation strategy together due to the lack of data.

Willingness to repeat cervical screening differed across the self-sampling device. *Supplemental Figure 10* shows that the willingness was higher for brushes and tampons compared with swabs (brush willingness 95% (95% CI: 90% to 97%) and tampon willingness 97% (95% CI: 92% to 99%) versus swab 87% (95% CI: 82% to 90%), with p = 0.007 for inclusion of sampling deviance as covariate). There was not sufficient data, or in a consistent format, for ethnicity or age to be considered in a quantitative manner.

## Discussion

In 2021, WHO published a guideline recommending the use of HPV DNA based screening strategies including HPV DNA self-sampling in place of pap smear and VIA screening. Our findings of HPV self-sampling support this strategy. The pooled absolute sensitivity of hrHPV assays for CIN2+ and CIN3+ were lower for self-sampling than for health professional sampling, for both colposcopy referral and primary screening. In contrast, the pooled absolute specificity of hrHPV assays for CIN2+ was greater for self-sampling than for health professional sampling for colposcopy referral, but not for primary screening. However, the difference was not statistically significant. The agreement between the screening test was 87.1% and a kappa value of 0.70. The pooled participation was higher in the mail-to-all self-sampling strategies compared to control. Cervical cancer screening non-attendees generally accept self-sampling (91%).

### Accuracy of HPV testing in self-collected samples compared with health professional-collected samples

Our findings are consistent with those reported in the source review(14). In high income countries, the interest in HPV DNA self-sampling has been for non/under screeners attendees (14). However, our review has included women of the general population with majority of studies including women who were referred for colposcopy or those attending the primary screening. In the UK, the HPValidate study found relative sensitivity and specificity of self-sampling and clinician collected samples to be high. Relative sensitivity ranging from 76.7% - 92.4% and specificity from 86.7% - 98.8% based on the different combinations of self-sampling devices and assays(30).

### The level of concordance between HPV-DNA testing in self-collected samples and health professional collected samples in cervical screening non-attenders

These findings are consistent with the findings from Arbyn et al 2022 which reported pooled estimates of agreement of 88.7% and the kappa of 0.72 (31). These results are still consistent with this global study which had a few studies from countries with higher burden of cervical cancer due to lack of well-established screening programs. The finding of this study was challenged on the fact that most studies included been a colposcopy referral population (19 studies), there was a lack of inclusion of studies from developing countries with no routine screening program and the use of kappa(32). The similarity maybe due to few studies which were included in the previous review from high-risk population (three studies). Similar to a previous review, this review had most study populations being referred to colposcopy (26/38). Hence there is less evidence from the self-sampling from primary screening. As new evidence are becoming available, a systematic review and meta-analysis of evidence emanating from primary screening by using self-sampling is warranted. In our subgroup analysis, the overall agreement was higher in the target amplification-based DNA assay compared to other assays. In Arbyn’s analysis, the test positivity ratio did not change between the signal amplification assay and target amplification assay(31). However, in this analysis, it was recommended that test positivity ratios may not be appropriate for predicting the clinical sensitivity of SA tests of self -vs clinician-collected samples(31). This is because the specificity of SA is lower with self-sampling, and the higher test sensitivity of the SA is associated with false positive results instead of true positive. The possible biological explanation of this is the lower load of HPV in the vagina and cross-reactions with low-risk HPV types with SA(31). The source review had many women who were referred for colposcopy. As new evidence become available, there is a need to carry out systematic reviews on this newly available evidence of self-sampling as a primary test.

### Uptake of cervical screening by HPV self-sampling method when compared to health professional sampling method in non-attenders with those offered health professional sampling

The findings of this review is consistent with the reference review(9). Mail to all have shown increased uptake, however, it may not be the optimal strategy when implementing self-sampling into clinical practice. Mail-to-all is costly and may result in significant wasted resources, as the majority do not return the kit. In a trial done in UK assessing a good screening strategy for women at their first invitation for cervical cancer screening found mail to all leading to better screening uptake and was more likely to be cost effective but at a higher cost per woman screened. In this study, the cost per woman screened under opt in strategy was £2.03 while it was £62.00 for the mail to all strategy(33).

The previous reviews recommended countries level pilots due to variation in the included studies(13,14). Due to this, countries including UK have done pilot studies for HPV self-sampling. In the UK the YouScreen study was a feasibility clinical trial embedded within the Cervical Screening Programme in England Programme to estimate the impact of offering self-sampling to non-attenders in practice(16). Self-sampling kits were offered opportunistically in-person in GP primary care and offered systematically via direct mailout. In the opportunistic offering of sampling arm, 65.5% returned self-samples compared with 12.9% in the systematically direct mailout arm(16). The study concluded that opportunistic offering of self-sampling could potentially increase uptake of cervical cancer screening to non-attendees and increase equity within HICs organised screening programmes. In this study, self-sampling was able to increase coverage for non-attendees by 22% per month. Our rapid review found one study that offered opportunistic self-sampling kit in GP primary care which it uptake was 6.9%(34), but our data on mail-to-all self-sampling reported similar participation rates (17.7% to 23%). YouScreen showed self-sampling resulted in a 22% increase and 12% increase in non-attenders screened per month from the per protocol and intention-to-treat analysis, respectively(16). Our meta-analysis of the literature also reported an increase in uptake, but the effect was more modest.

We found a low percentage of unsatisfactory samples while adherence to follow-up was higher encouraging the applicability of this method. Whilst the small percentage of the unsatisfactory sample may be reassuring for those whom doubts regarding their self-efficacy in performing self-sampling is a barrier to participation, this is likely an under-estimate and may vary according to the device utilised(35). One of the challenges of self-sampling is loss of follow-up, however, this level of adherence assures the linkage of those with positive results to further assessment for identification of precancer and cancer.

Despite including only studies from HICS, heterogeneity was still present limiting generalizability of the findings. Therefore, calling for country specific feasibility studies including also cost effectiveness, logistics and compliance studies.

### Acceptability of HPV self-sampling screening strategies to those that have not attended the regular cervical screening programme

We found that, self-sampling is acceptable to non-attendees. In the context of the UK, the HPValidate study found self-sampling to be acceptable(36,37). In the HPValidate study, 69% of participants preferred self-sampling at home(36) and over half of participants (54.6%) preferred the self-test over clinician collected samples(37). This finding shows that, acceptability of these methods can increase uptake, reduce disparities in screening and cervical cancer burden in HICs in which studies have shown it most affect women who do not attend in the routine screening. The reference review reported pooled reasons for preferring self-sampling were ease of use (91%), not embarrassing (91%), privacy (88%), comfort performing self-sampling (88%), ability to do it oneself (69%) and convenience (65%) (13). The most reported pooled reason for disliking was the uncertainty of doing it correctly (21%), pain or physical uncomfortable (10%), anxiety (15%) and not wanting to touch themselves (6%) (13). Our meta-analysis found that self-sampling led to pain or discomfort (18%), caused embarrassment (12%), caused anxiety (35%) and did not fit with their values (60%). Despite these reported advantages of self-sampling, the strategy may exclude those with disabilities which limit their ability to self-sample such as people with visual impairments, motor dysfunction, or mental health issues.

Data was limited regarding reasons for liking or disliking self-sampling for non-attenders. Indeed, the data is available for studies that were newly extracted but were not available for the studies in the existing review. Most of the studies included were cross sectional surveys and performed poorly on the risk of the assessment. 46/48 studies scores 4 points or below on the eight items scores. Most of the studies did not report on the items in the assessment tool. Because of the cross sectional design of the included study, the causal inference is not drawn and is not possible to tell if this will be applicable in a non-study environment(13). Like the previous global review, we also observed high heterogeneity which father limits the generalizability of the findings.

### Strength and Limitations

This is a comprehensive rapid review of the existing literature in HPV self-sampling for cervical cancer screening. Rather than focusing on an aspect of self-sampling in isolation e.g acceptability, this review gives a more holistic understanding on the usefulness of HPV self-sampling in population screening programmes for cervical cancer. It only includes developed countries which are more advanced in cervical screening compared to developing countries. However, there are limitations to this analysis. Some studies were not included in the meta-analysis as data requirements not fulfilled from the existing reviews. Due to the rapid nature of the review, many study results were extracted from existing reviews. Unfortunately, only the relative sensitivity and specificities were reported in the review(s) we utilized for the accuracy question, which could not be used to back-calculate the necessary 2×2 tables, and so were excluded from the relevant meta-analyses. Secondly, the statistical methods used to calculate the pooled estimate for the accuracy question do not consider the ‘paired’ nature of the studies (i.e. the fact that it was the same women in the ‘self’ and the ‘health’ arms for each study). However, we believe that the consequence, if there is any, of not taking this into account means the estimates above (95% CI) may be slightly conservative. Most of the studies in the reference reviews for concordance and accuracy, the population were for women who were referral for colposcopy. Finally, the assessment of subgroups was not possible due to limited data from the studies from the reference review and the study not analysing the outcome at the subgroup level. There was not sufficient data, or a consistent format, for ethnicity or age to be considered quantitatively.

Participation is often reduced in some patient populations, including those in minority ethnic groups, those of low socio-economic status, and transgender and non-binary people with a cervix, but there were insufficient data to explore the possible impact of self-sampling in these populations in our review. We have reported four ongoing studies, non – of this also has considered these subgroups(38–41). These studies have focused on the role of HPV self-sampling to clinician collected samples on increasing uptake of cervical cancer screening for non-attendees. One of this study compared the uptake of HPV self-sampling with the self-sampling devices offered by the GP and the kits been sent mailed(39).

### Conclusion

Self-sampling is a feasible strategy for reaching non-attendees in and should be considered in the national screening program to reach the non-attendees, especially on using the PCR-based assay. However, before this is done, understanding the cost-effectiveness, logistics and compliance of the strategies is important to understand country-specific strategies for reaching the non-attendees.

## Sources

This rapid review was conducted by the Evidence Synthesis Group at the Complex Reviews Synthesis Unit (ESG @CRSU). Evidence relevant to the most recently published YouScreen study, which estimated the impact of offering HPV self-sampling to non-attenders within the cervical screening programme in England, was synthesised(16).

## Funder

This study was funded by the NIHR Evidence Synthesis Programme.

## Authors’ contribution

NT: Screening of the articles, data extraction, manuscript writing and coordination

CN: Data analysis and writing of the manuscript

MT: Data extraction and manuscript writing

RM: Conceptualization, developed search strategy, screening of the articles, data extraction, manuscript writing

AN: Conceptualization, development of search strategy and querying the database

TQ: Supervision, reading and editing of the manuscript

AS: Supervision, reading and editing of the manuscript. Also, contributed to study design and analysis method.

OW: Supervision, reading and editing of the manuscript

## Data Availability

All data produced in the present work are contained in the manuscript supplimentary material

