## Supplementary material for "HPV Self-Sampling in Cervical Screening: A Rapid Review": Suplimental Material

**Title: HPV Self-Sampling in Cervical Screening: a Rapid Review: Supplemental Materials**

### Supplementary
### Methodological Approaches for each research question (Include PICOS, bibliographic databases and analysis)

What is the accuracy of HPV testing in self-collected samples compared with health professional collected samples, and does this vary according to eligible women and test characteristics?

A prior review by Arbyn et al was used as a basis in addressing this question(1)

<table>
<tr><td>Population</td><td colspan="3">Individuals eligible for cervical screening*</td></tr>
<tr><td>Index Test</td><td colspan="3">HPV testing on self-collected sample</td></tr>
<tr><td>Comparator Test</td><td colspan="3">HPV testing on healthcare professional-collected sample</td></tr>
<tr><td>Reference Standard</td><td colspan="3">Colposcopy +/- biopsy as indicated</td></tr>
<tr><td>Co-variates (where available)</td><td colspan="3">• Background risk of population<br>• Screening history of population (e.g under-screened, never screened)<br>• Clinical history of population (e.g HIV positive)<br>• Testing methodology<br>• Sampling method/kit<br>• Storage medium<br>• Home-based vs in-clinic self-sampling<br>• Age; Socioeconomic status; Ethnicity</td></tr>
<tr><td>Outcomes (where available)</td><td colspan="3">• Absolute sensitivity and specificity of HPV self-sampling for the detection of CIN2+ and CIN3+ of index and comparator tests<br>• Relative sensitivity and specificity of HPV self-sampling for CIN2+ and CIN3+ of HPV self-sampling versus clinician-based sampling<br>• False-positive and false-negative rates of HPV self-sampling versus clinician-based sampling<br>• PPV and NPV of HPV self-sampling<br>• Proportion of self-selected samples in which HPV status cannot be determined (e.g. insufficient sample, failed lab tests)<br>• Proportion of women with a ‘failed’ test/sample who are asked to provide a second sample<br>• Proportion of women with a positive test result who attend clinic for diagnostic investigations and treatment (including cytology follow-up)</td></tr>
<tr><td>Study designs</td><td colspan="3">Cross-sectional studies, cohort studies, randomised controlled trials (RCTs), systematic reviews.</td></tr>
<tr><td>Electronic databases</td><td>Database:<br>☒ MEDLINE<br>☒ CENTRAL<br>☒ EMBASE<br>☒ Clinical Trial Registry (ClinicalTrials.gov)</td><td>From:<br>1st January 2018 (overlap with Arbyn et al. 2018)</td><td>To:<br>March 2024</td></tr>
</table>

**These include “women who were irregularly screened, never screened, or did not respond to invitation or reminder letters for conventional screening for cervical cancer”, as defined in the Abryn review.*

Analyses were conducted according to the methods recommended in the Cochrane Handbook for Systematic Review of Diagnostic Test Accuracy (utilising the supplementary material in Chapter

10)(2). For each study, 2x2 tables for self-sampling (self) and healthcare professional sampling (health) were either extracted or backcalculated from the absolute sensitivities and specificities (with variance calculated from 95% confidence intervals) for self and health. Using the {lme4}(3) package, a single model was defined that included both sensitivity and specificity for self and health, together, with separate variances for self and health. This model gave pooled estimates of absolute sensitivity and specificity for self and health. Using the {msm}(4) package, the pooled absolute and relative difference between self and health for sensitivity and specificity could be calculated using the delta method(5) for calculating the confidence intervals. Absolute and relative differences were estimated separately for screening and colposcopy referral populations, and CIN2+ and CIN3+. Assay testing methodology and self-sampling device and setting were tested regarding affecting the outcome. These were tested by adding them to the model and then comparing models using the likelihood ratio test.

To calculate the pooled estimates appropriately (as per the Cochrane Handbook), the raw 2x2 data table are required. The reference reviews only reported relative sensitivity and specificity for each study, which was not sufficient to back-calculate the requisite data. Furthermore, of all the studies identified from the top-up search, eight did not have the necessary data (e.g. no comparator; no standard error). This has reduced the number of studies available for meta-analysis to 13 studies.

*The level of concordance between HPV-DNA testing in self-collected samples and health professional collected samples in cervical screening non-attenders*

A prior review by Arbyn et al was used as a basis in addressing this question, with specific additional consideration of an updated review and meta-analysis on concordance between self-collected and clinician-collected samples for HPV testing(1,6).

| | | | |
|---|---|---|---|
| **Population** | Individuals eligible for cervical screening | | |
| **Index test** | HPV testing on self-collected specimens | | |
| **Comparator/reference standard** | HPV testing on healthcare professional-collected specimens in index test subject | | |
| **Co-variates (where available)** | • Background risk of population<br>• Clinical history of population<br>• Testing methodology<br>• Sampling method/kit<br>• Storage medium<br>• Home-based vs in-clinic self-sampling<br>• Age; Socioeconomic status; Ethnicity<br>• Comorbidities captured by clinical history | | |
| **Outcomes (where available)** | • HPV status<br>• Test positivity ratio<br>• Percent positive agreement<br>• Percent negative agreement<br>• Cohen's Kappa statistic<br>• Positive concordance<br>• Negative concordance | | |
| **Study designs** | RCTs, cohort studies, systematic reviews | | |
| **Electronic databases** | Database: | From: | To: |

| | | | |
|---|---|---|---|
| | ☒ MEDLINE<br><br>☒ CENTRAL<br><br>☒ EMBASE<br><br>☒ Clinical Trial Registry (ClinicalTrials.gov) | 1st January 2018 (overlap with Arbyn et al. 2018) | March 2024 |

Test positivity rate ratio, overall agreement, positive agreement, negative agreement, kappa, positive concordance, and negative concordance were meta-analysed – note that all of these measures were extracted concordant outcomes from the studies and were comparing self-samples vs healthcare professional collected results. Test positivity rate ratio was meta-analysed with {metafor} using a log transformation. Kappa was meta-analysed with {metafor} and utilised the measure of overall agreement to estimate variance when applicable(7). The remaining outcomes were meta-analyses of proportions using the metaprop command in {meta}. Assay testing methodology, self-sampling setting, and self-sampling device were tested regarding influencing the outcomes.

*Uptake of cervical screening by HPV self-sampling method when compared to health professional sampling method in non-attenders with those offered health professional sampling*

A prior review by Arbyn et al was used as a basis in addressing this question(1).

| | |
|---|---|
| **Population** | Individuals who were invited to participate in standard cervical screening programme but did not respond to invitation or did not participate in the screening programme |
| **Intervention** | Invitation to HPV based cervical screening - self sampling: opt-in, mailed, door-to-door, opportunistic |
| **Comparator** | Invitation to HPV based cervical screening - clinician/health professional sampling |
| **Co-variates (where available)** | • Invitation strategy (including opt-in; opt-out; opportunistic)<br>• Screening history<br>• Time from invitation for clinician/health professional sampling<br>• Clinical history of population<br>• Sampling method (brush, swab, lavage)<br>• Location of test (home vs clinic/primary care)<br>• Use of reminders (e.g. SMS)<br>• Age; Socioeconomic status; Ethnicity<br>• Comorbidities |
| **Outcomes (where available)** | • Uptake of HPV based cervical screening (absolute participation)<br>• Relative participation<br>• Participation difference<br>• Adherence to follow-up among those with a positive test result<br>• PPV for CIN2+ among those with a positive test that attended for follow-up |

| | | | |
|---|---|---|---|
| | • Proportion of self-sampling individuals with unsatisfactory test results, i.e HPV status cannot be determined (e.g. insufficient sample, failed lab tests)<br>• Proportion of women with a 'failed' test/sample who are asked to provide a second sample<br>• CIN2+ detection rate<br>• Frequency of screening across rounds | | |
| **Study designs** | RCTs, cohort studies, systematic reviews | | |
| **Electronic databases** | Database:<br>☒ MEDLINE<br>☒ CENTRAL<br>☒ EMBASE<br>☒ Clinical Trial Registry (ClinicalTrials.gov) | From:<br>1st January 2018 (overlap with Arbyn et al. 2018) | To:<br>March 2024 |

Within each study, absolute participation was defined as the number of responders divided by the total number of individuals invited for the respective screening technique. Participation difference was then defined as the difference between these absolute percentages (i.e. self-sampling – control), and relative participation was calculated by dividing the absolute percentage of responders in the self-sampling group by the absolute percentage of responders in the control group. Absolute participation (self-sampling and control), unsatisfactory sample, adherence to follow-up, and CIN2+ detection were pooled using the metaprop command in {meta}. Participation difference and relative participation were meta-analysed using the metabin command in {meta}. Absolute and relative participation outcomes were meta-analysed separately for per protocol/intention-to-treat analysis results and invitation scenario. Per protocol analysis included women who participated in the cervical cancer screening through an HPV DNA self-sampling arm only. Intention-to-treat analysis included also those who were invited for self-sampling but chose to have a clinician-collected sample instead. Self-sampling device, whether reminders were used, and time between invitation and healthcare professional sampling were tested regarding influencing the outcomes,

*Acceptability of HPV self-sampling screening strategies to those that have not attended the regular cervical screening programme*

A prior review by Nelson et al was utilized as the basis for addressing this question, with particular consideration of additional reviews by Yeh et al and Nishimura et al(8,9)

| | |
|---|---|
| **Population** | Individuals eligible for cervical screening who do not attend for health professional testing |
| **Intervention** | Invitation to HPV-based cervical screening - self-sampling |
| **Comparator** | Invitation to HPV-based cervical screening - health professional sampling |

| | | | |
|---|---|---|---|
| **Co-variates (where available)** | • Invitation strategy<br>• Sampling method (brush, swab, lavage)<br>• Screening history<br>• Clinical history of population<br>• Population subgroup (eg SES, ethnicity, LGBT+) | | |
| **Outcomes (where available)** | Overall:<br>• Stated overall acceptability<br>• Stated preference in compared with clinician-based screening<br>• Stated preference for the setting of self-collection of sample<br>• Stated willingness to repeat screening<br>Individual characteristics of acceptability/experience including:<br>• Logistic measures of acceptability (e.g convenience, accessibility)<br>• Procedure-related measures of acceptability (e.g pain/physical discomfort, ease of use, confidence in result, self-efficacy to do the test)<br>• Psychosocial measures of acceptability (e.g stigma, embarrassment, anxiety, fit with values) | | |
| **Study designs** | RCTs, cohort studies, feasibility studies, mixed methods studies, surveys and systematic reviews. | | |
| **Electronic databases** | Database:<br>☒ MEDLINE<br>☐ CENTRAL<br>☒ EMBASE<br>☒ Other (CINAHL, LILACS, SCOPUS, OpenGrey, ProQuest, Cochrane Library)<br>☒ Clinical Trial Registry (ClinicalTrials.gov) | From:<br>1st December 2014 (overlap with Nelson et al. 2015) | To:<br>March 2024 |

All outcomes were meta-analysed using the metaprop command in {meta}. Due to data availability, only self-sampling devices were tested regarding influencing the outcomes.

### Statistical Analysis

The analysis for all questions followed the Cochrane Handbook for Systematic Review of Diagnostic Test Accuracy where appropriate:

- Meta-analyses were primarily conducted in R(10) using the {meta}(11)or {metafor} package(12). Where necessary, the variance for each study was estimated from the reported confidence intervals using the conv.wald command in {metafor}.
- Forest plots were produced to investigate potential heterogeneity in meta-analyses. For each forest plot, studies were ordered by year to assess any temporal patterns.

- Outcomes were pooled separately by characteristics that were known to give inherently different results.
- Where data permitted, meta-regressions were conducted to assess whether certain characteristics influenced outcomes and explained any between-study heterogeneity. Characteristics were considered in separate regression models. Where a characteristic had a p-value $< 0.05$, a respective subgroup forest plot was produced.

Supplemental Table I: Search Strategies

*Clinical Accuracy (per Arbyn et al.)*(1)

| Database | Search |
|---|---|
| PubMed | #1: Cervix OR cervico* OR cervica*<br>#2: Cancer OR carcinoma OR neoplas* OR dysplas* OR CIN[tw] OR CINII*[tw] OR CIN2*[tw] OR CINIII*[tw] OR CIN3[tw] OR SIL[tw] OR SIL OR HSIL[tw] OR H-SIL OR LSIL[tw] OR L-SIL OR OR ‘‘low grade’’ OR low-grade OR mild OR equivocal OR borderline.<br>#3: #1 AND #2.<br>#4: HPV OR "Human Papillomavirus DNA Tests"[Mesh] OR ‘‘human papillomavirus’’ OR papillomavir* OR viral OR virus<br>#5: self-collection OR “self collection” OR self-sampling OR self-collect* OR self-sampl* OR self OR "Self- Examination"[Mesh]<br>#6: #4 AND #5<br>#7: #3 AND #6<br>#8: Publication Date from January 2018 to March 2024.<br>#9: #7 AND #8 |
| Embase | #1: 'cervix'/exp OR cervix OR cervico* OR cervica*<br>#2: 'cancer'/exp OR cancer OR 'carcinoma'/exp OR carcinoma OR neoplas* OR dysplas* OR cin OR 'cin2' OR 'cin3' OR sil OR hsil OR h+sil OR lsil OR l+sil OR 'low grade' OR low+grade OR mild OR equivocal OR 'borderline'/exp OR borderline<br>#3: 'hpv'/exp OR hpv OR 'human papillomavirus'/exp OR 'human papillomavirus' OR papillomavir* OR viral OR 'virus'/exp OR virus<br>#4: self+collection OR 'self collection' OR self+sampling OR 'self-sampling' OR self+collect* OR self+sampl* OR 'self'/exp OR self<br>#5: #1 AND #2 AND #3 AND #4<br>With the following limits:<br>• - Map to preferred terminology (with spell check)<br>• - Also search as free text<br>• - Include sub-terms/derivatives (explosion search) |
| Cochrane Library | #1: Cervix or cervico* or cervica*<br>#2: Cancer or carcinoma or neoplas* or dysplas* or CIN or CIN2 or CIN3 or SIL or SIL or HSIL or H-SIL or LSIL or L-SIL or "low grade" or low-grade or mild or equivocal or borderline.<br>#3: HPV or ‘‘human papillomavirus’’ or |

| | |
|---|---|
| | papillomavir* or viral or virus<br>#4: self-collection or "self collection" or self-sampling or ‘‘self-sampling’’ or self-collect* or self-sampl* or self<br>With the following limits:<br>• Cochrane reviews (reviews + protocols)<br>• Other reviews<br>• Search for word variations |

*Strategies to increase population coverage of cervical screening (Albyn et al.)*(1)

| **Database** | **Search** |
|---|---|
| PubMed | (Cervix OR cervical) AND (HPV OR papillomavirus) AND (self-sampling OR self sampling OR self-collection OR self collection) AND (screening OR coverage OR participation OR knowledge OR acceptance) |

*Acceptability*
(per Nelson et al)(8)

| **Database** | **Search** |
|---|---|
| ProQuest Dissertations and Theses | (Prefer* OR feasib* OR accept* OR barrier OR cost OR attitude) AND (HPV OR "Human papillomavirus") AND (self-collect* OR self-sampl* OR self-screen*) |
| PubMed | (("human papillomavirus"[All Fields] OR HPV[All Fields]) AND (accept[All Fields] OR prefer[All Fields] OR ("attitude"[MeSH Terms] OR "attitude"[All Fields]) OR barrier[All Fields] OR fesi[All Fields] OR ("economics"[Subheading] OR "economics"[All Fields] OR "cost"[All Fields] OR "costs and cost analysis"[MeSH Terms] OR ("costs"[All Fields] AND "cost"[All Fields] AND "analysis"[All Fields]) OR "costs and cost analysis"[All Fields]))) AND (self-collection[All Fields] OR self-collect[All Fields] OR self- sampling[All Fields] OR self-sample[All Fields] OR self-screen[All Fields]) |
| SCOPUS | (TITLE-ABS-KEY ( "human papillomavirus" OR hpv ) AND TITLE-ABS-KEY ( accept OR prefer OR attitude OR barrier OR feasib OR cost ) AND TITLE- ABS-KEY ( self-collection OR self-collect OR self-sampling OR self-sample OR self-screen ) ) |
| Web of Science | TOPIC: ("human papillomavirus" OR HPV) AND TOPIC: (accept OR prefer OR attitude OR barrier OR cost OR feasib) AND TOPIC: (self-collection OR self-collect OR self-sampling OR self-sample OR self-screen) Timespan: All years. Indexes: SCI-EXPANDED, SSCI, A&HCI. |

| | |
|---|---|
| OpenGrey | (HPV OR "Human papillomavirus") AND (collect* OR Sampl* OR screen*) HPV OR "Human papillomavirus" |
| Cochrane Database of Systematic Reviews | HPV OR "Human papillomavirus" |

(*per* Yeh et al. and Nishimura et al)(9,13)

| **Database** | **Search** |
|---|---|
| PubMed | ("human papillomavirus"[tiab] OR HPV[tiab] OR "cervical"[tiab] OR "cervix"[tiab]) AND ("self-test" [tiab] OR "self-testing" [tiab] OR "home-based test"[tiab] OR "home-based testing"[tiab] OR "home test"[tiab] OR "home testing"[tiab] OR "clinic-based test"[tiab] OR "clinic-based testing"[tiab] OR "community-based test"[tiab] OR "pharmacy-based test"[tiab] OR "self-administer"[tiab] OR "self-sampling"[tiab] OR "self-collecting"[tiab] OR "self-collected"[tiab] OR "self-collection"[tiab] OR "self- versus provider-collected"[tiab] OR "self- and provider-collected"[tiab] OR "self- versus physician- collected"[tiab] OR "self- and physician-collected"[tiab] OR "self care"[Mesh] OR self- administration[Mesh] OR "self assessment"[Mesh]) |
| CINAHL | (TI "human papillomavirus" OR TI HPV OR TI cervical OR TI cervix OR AB "human papillomavirus" OR AB HPV OR AB cervical OR AB cervix) AND (TI “self-test” OR AB “self-test” OR TI "self-testing" OR AB “self-testing” OR TI “home-based test" OR AB “home-based test” OR TI "home-based testing" OR AB “home-based testing” OR TI "home test" OR AB “home test” OR TI "home testing" OR AB “home testing” OR TI "clinic-based test" OR AB “clinic-based test” OR TI "clinic-based testing" OR AB “clinic-based testing” OR TI "community-based test" OR AB “community-based test” OR TI "pharmacy-based test" OR AB “pharmacy-based test” OR TI "self- administer" OR AB “self-administer” OR TI "self-sampled" OR AB “self-sampled” OR TI "self-sample" OR AB “self-sample” OR TI "self-sampling" OR AB “self-sampling” OR TI "self-collecting" OR AB “self- collecting” OR TI "self-collected" OR AB “self-collected” OR TI "self-collection" OR AB “self-collection” OR TI "self- versus provider-collected" OR AB “self- versus provider-collected” OR TI "self- and provider- collected" OR AB “self- and provider-collected” OR TI "self- versus physician-collected" OR AB “self- |

| | |
|---|---|
| | versus physician-collected" OR TI "self- and physician-collected" OR AB "self- and physician-collected") |
| Embase | ('human papillomavirus':ab,ti OR HPV:ab,ti OR cervical:ab,ti OR cervix:ab,ti)<br>AND<br>('self-test':ab,ti OR 'self-testing':ab,ti OR 'home-based test':ab,ti OR 'home-based testing':ab,ti OR 'home test':ab,ti OR 'home testing':ab,ti OR 'clinic-based test':ab,ti OR 'clinic-based testing':ab,ti OR 'community-based test':ab,ti OR 'pharmacy-based test':ab,ti OR 'self-administer':ab,ti OR 'self- sampled':ab,ti OR 'self-sample':ab,ti OR 'self-sampling':ab,ti OR 'self-collecting':ab,ti OR 'self- collected':ab,ti OR 'self-collection':ab,ti OR 'self- versus provider-collected':ab,ti OR 'self- and provider-collected':ab,ti OR 'self- versus physician-collected':ab,ti OR 'self- and physician-collected':ab,ti) |
| LILACS | ("human papillomavirus" OR HPV OR cervical OR cervix) [words]<br>AND<br>("self-test" OR "self-testing" OR "home-based test" OR "home-based testing" OR "home test" OR "home testing" OR "clinic-based test" OR "clinic-based testing" OR "community-based test" OR "pharmacy-based test" OR "self-administer" OR "self-sampling" OR "self-collecting" OR "self-collected" OR "self-collection" OR "self- versus provider-collected" OR "self- and provider-collected" OR "self-versus physician-collected" OR "self- and physician-collected") [words] |

### Supplemental Table 2: Characteristics of Studies on Test Accuracy of HPV Testing in Self-selected Samples

| Author, Year and Country | Population | Sample Size | Age (years) | Ethnicity | Device used | Setting | hrHPV Assay | Storage Medium | Outcomes Assessed |
|---|---|---|---|---|---|---|---|---|---|
| Hillemanns 1999 Germany(14) | Colposcopy referral | 247 | Not specified | No reported | Brush | Not specified | HC2 | "placed into a specimen collection tube" | CIN2+ |
| Sellors 2000 Canada(15) | Colposcopy referral | 200 | Mean 31.5 Range not given | Not reported | Swab | Not specified | HC2, PCR (L1 consensus) | Self: STM Clin brush: STM Clin swab: sterile phosphate buffered saline | CIN2+ |
| Nobbenhuis 2002 The Netherlands(16) | Colposcopy referral | 71 | Mean 35 Range not given | Not reported | Lavage | Not specified | PCR | PBS | CIN2+ |
| Brink 2006 The Netherlands(17) | Colposcopy referral | 96 | Median 35 Range 18-59 | Not reported | Lavage | Not specified | PCR | SurePath | CIN2+ |

| | | | | | | | | | |
|---|---|---|---|---|---|---|---|---|---|
| Szarewski 2007<br><br>UK(18) | Primary screening | 920 | Median 29 (population 1)<br>Median 41 (population 2) | Not reported | Swab | Not specified | Not specified | | CIN2+ |
| Balasubramanian 2010<br><br>USA(19) | Primary screening (high risk) | 1665 | Median 23<br>Range 18-50 | Not reported | Swab | Not specified | HC2 | STM | CIN2+ |
| Dijkstra 2012<br>The Netherlands(20) | Colposcopy referral | 135 | Median 34<br>Range not given | Not reported | Brush | Not specified | PCR | Cell preserving | CIN2+ |
| van Baars 2012<br><br>The Netherlands(21) | Colposcopy referral | 134 | Mean 40<br>Range 21-66 | Not reported | Brush | Not specified | PCR | Self: FTA cartridge<br>Clin: ThinPrep, SurePath | CIN2+<br>CIN3+ |
| Jentschke 2013a<br><br>Germany(22) | Colposcopy referral | 72 | Mean 37<br>Range 16-68 | Not reported | Lavage | Not specified | HC2 | Self: buffered saline<br>Clin: PreservCyt, Cervatec | CIN2+<br>CIN3+ |
| Jentschke 2013b<br><br>Germany(23) | Colposcopy referral | 42 | Mean: 36<br>Range: 18-68 | No reported | Lavage | Not specified | hrHPV: HC2 | Self: buffered saline | CIN2+<br>CIN3+ |

| | | | | | | | | | |
|---|---|---|---|---|---|---|---|---|---|
| | | | | | | | P16: p16INK4a ELISA | Clin: PreservCyt, Cervatec | |
| Stanczuk 2016<br><br>UK(24) | Primary Screening | 5318 | Mean 41<br>Range 18–76 | Not reported | Swab | Not specified | PCR | Cell preserving | CIN2+<br>CIN3+ |
| Jentschke 2016<br><br>Germany(25) | Colposcopy referral | 136 | Mean 36<br>Range 17–78 | Not reported | Brush | Not specified | PCR | Cell preserving | CIN2+<br>CIN3+ |
| Aiko 2017<br>Japan(26) | Colposcopy referral | 136 | Mean not given<br>Range 20-69 | Not reported | Brush | Not specified | HC2 | | CIN2+<br>CIN3+ |
| Asciutto 2017<br><br>Sweden(27) | Colposcopy referral | 218 | Mean 35<br>Range 19-71 | Not reported | Swab | Not specified | PCR | Cell preserving | CIN2+ |
| Leeman 2017<br><br>The Netherlands(28) | Colposcopy referral | 91 | Mean not reported<br>Range 18-60 | Not reported | Brush | Not specified | PCR | Cell preserving | CIN2+<br>CIN3+ |
| Catarino 2017<br><br>Switzerland(29) | Colposcopy referral | 150 | Median 32<br>Range 18-69 | Not reported | Swab | Not specified | PCR | Cell preserving | CIN2+<br>CIN3+ |

| | | | | | | | | | |
|---|---|---|---|---|---|---|---|---|---|
| Leinonen 2018<br><br>Norway(30) | Patients with cervical premalignant lesion and carcinoma diagnosis or carcinoma suspicion | 240 | Mean 38<br><br>Range 21-80 | Not reported | Brush and Swab | Not specified | PCR | Cell preserving | CIN3+ |
| Leinonen 2018<br><br>Norway(30) | Colposcopy referral | Self sampling:<br><br>Evalyn Brush=287;<br><br>FLOQ swabs=286<br><br>Health professional sampling: 259 | Not reported | Not reported | Swab and Brush | Home | PCR | Cell preserving | CIN3+ |
| Igidbashian 2014<br><br>Italy(31) | Primary screening | 700 | Mean 44.3<br><br>Range. Not reported | Not reported | Brush | Clinical setting | The Hybrid Capture II (HC2) microplate method | Cell preserving | CIN2+ |
| Mangold 2019 | Colposcopy referral | 208 | Not reported | Not reported | Swab | Not specified | Signal amplification and PCR | Cell preserving | CIN2+ |

| | | | | | | | | | |
|---|---|---|---|---|---|---|---|---|---|
| Germany(32) | | | | | | | | | |
| Edbald-Svensson 2018<br><br>Sweden(33)* | Colposcopy referral | 63 | Mean 42<br>Range 24–64 | Not reported | Swab | Not specified | PCR | Cell preserving | CIN2+ |
| El-Zein 2018<br><br>Canada(34)* | Colposcopy referral | 1217 | Not reported | Not reported | Swab | Clinical setting | PCR | Cell preserving | CIN2+<br>CIN3+ |
| El-Zein 2019<br><br>Canada(35) | Colposcopy referral | 700 | Mean 37.7<br>Range not reported | Not reported | Swab | Clinical setting | PCR | Cell preserving | CIN2+<br>CIN3+ |
| Polman 2019<br><br>The Netherlands(36)* | Primary screening | Self sampling: 7643<br>Health professional sampling: 6282 | Self sampling mean= 45·5<br>Clinician based sampling mean = 45·7<br>Range not given | No reported | Brush | Home | PCR | Cell preserving | CIN2+<br>CIN3+ |
| Onuma 2020<br><br>Japan(37)* | (1) Outpatients with abnormal cytology and requiring colposcopy | 100 | Mean 41.8<br>Range not given | Not reported | Brush | Clinical setting | Cobas 4800 system (PCR) | Cell preserving | CIN2+ |

| | | | | | | | | | |
|---|---|---|---|---|---|---|---|---|---|
| | and biopsy and (2) NILM/HPV-positive patients in the Fukui Cervical Cancer Study | | | | | | | | |
| Ørnskov 2020<br><br>Denmark(38)* | Colposcopy referral | 305 | Median 34<br><br>Range 17-85 | Not reported | Brush | Clinical setting | PCR | Cell preserving | CIN2+<br><br>CIN3+ |
| Cho 2020<br><br>South Korea(39)* | Colposcopy referral | 314 | Median 40<br><br>Range not reported | Not reported | Brush | Clinical setting | PCR | Cell preserving | CIN2+ |
| Rohner 2020a<br><br>USA(40) | Colposcopy referral | 314 | Median 36<br><br>Range not given | Non-Hispanic white: 38%<br><br>Hispanic: 29%<br><br>non-Hispanic Black: 26%<br><br>Other racial | Brush | Clinical setting | PCR | Cell preserving | CIN2+ |

| | | | | | | | | | |
|---|---|---|---|---|---|---|---|---|---|
| | | | | identities: 6% | | | | | |
| Rohner 2020b<br>USA(41)* | Colposcopy referral | 307 | Median 36<br>Range not given | Hispanic: 29%<br>Non-Hispanic white:38%<br>Non-Hispanic black:<br>26%; Other: 7% | Brush | Clinical setting | PCR | Cell preserving | CIN2+ |
| Ertik 2021<br>Germany(14)* | Colposcopy referral | 65 | Median age 36<br>Range 24–76 | Not reported | Swab and Brush | Home | PCR | Cell preserving | CIN2+ |
| Klischke 2021<br>Germany(42)* | Colposcopy referral | 70 | Mean 37 | Not reported | Brush | Clinical setting | PCR | Cell preserving | CIN2+<br>CIN3+ |
| Latsuzbaia 2022a<br>Belgium(43)* | Colposcopy referral | 485 | Median 40<br>Range not reported | No reported | Brush | Clinical setting | PCR | Cell preserving | CIN2+<br>CIN3+ |
| Avian 2022<br>Italy(44)* | Primary screening | 889 | Mean not reported | Not reported | Swab | Clinical setting | PCR | Cell preserving | CIN2+ |

| | | | | | | | | | |
|---|---|---|---|---|---|---|---|---|---|
| | | | 30-39: 190 (21.4%); 40-49: 303 (34.1%); 50-59: 299 (33.6%); ≥ 60: 97 (10.9%) | | | | | | |
| Latsuzbaia 2022b Belgium(45) | Colposcopy referral | 486 | Median 40 Range not reported | Not reported | Swab and Brush | Clinical setting | PCR | Cell preserving | CIN2+ CIN3+ |
| Stanczuk 2022 UK(46) | Primary screening | 4617 | Mean 41.3 Range not given | Not reported | Not specified | Not specified | Cobas 4800 PCR-based DNA test | ThinPrep (PreservCyt Solution Hologic, UK) | CIN2+CIN3+ |
| Latsuzbaia 2023a Belgium(47) | Colposcopy referral | 483 | Median 40 Range not reported | Not reported | Swab and Brush | Clinical setting | PCR and signal amplification | Cell preserving | CIN2+ CIN3+ |
| Latsuzbaia 2023b Belgium(48) | Colposcopy referral | 493 | Not reported | Not reported | Swab and Brush | Clinical setting | PCR | Cell preserving | CIN2+ CIN3+ |
| Martinelli 2023 Italy(49)* | Colposcopy referral | 245 | Median 38 Range not reported | Not reported | Swab | Clinical setting | PCR | Cell preserving | CIN2+ |

| | | | | | | | | | |
|---|---|---|---|---|---|---|---|---|---|
| Martinelli 2024<br><br>Italy(50)* | Colposcopy referral | 290 | Median 40<br><br>Range not reported | Not reported | Swab | Clinical setting | PCR | BD HPV Self Collection Diluent | CIN2+<br>CIN3+ |

Coloured red: From the top up search
* Indicate studies included in meta-analysis

**Supplemental Table 3: Studies with no outcome on Concordance between HPV-DNA Testing in Self and Health Professional Collected Samples from the review**

| Author, Year, Country | Population | Sample Size | Age (years) | Ethnicity | Device | Setting | hrHPV Assay | Storage medium |
|---|---|---|---|---|---|---|---|---|
| Sellors 2000<br><br>Canada(15) | Not reported | 200 | Mean 31.5 Range not given | Not specified | Swab | Clinical setting | Both HC2, PCR (L1 consensus) | Self: STM Clin brush: STM Clin swab: sterile phosphate- buffered saline |
| Daponte 2006<br><br>Greece(51) | Not reported | 98 | Not specified | Not specified | Brush | Clinical setting | PCR | PBS |
| Szarewski 2007<br><br>UK(18) | Not reported | 920 | Median 29 (pop 1) Median 41 (pop 2) | Not specified | Swab | Not specified | HC2 | Not specified |
| Balasubramanian 2010<br><br>USA(19) | High risk | 1665 | Median 23 Range 18-50 | Not specified | Swab | Not specified | HC2 | STM |
| Gustavsson 2011<br><br>Sweden(52) | Not reported | 50 | Mean not reported Range 39-60 | Not specified | Brush | Clinical setting | PCR | FTA cartridge |
| Twu 2011<br><br>Taiwan(53) | Unscreened for ≥3years | 252 | Median 42 Range 26-79 | Not specified | Brush | Clinical setting | PCR | STM |

| | | | | | | | | |
|---|---|---|---|---|---|---|---|---|
| Dijkstra 2012<br><br>The Netherlands(20) | Not reported | 135 | Median 34<br>Range not given | Not specified | Brush | Clinical setting | PCR | PreservCyt |
| Geraets 2013<br><br>Spain(54) | Not reported | 182 | Median 34<br>Range: 16-76 | Not specified | Brush | Clinical setting | PCR | FTA cartridge |
| Stanczuk 2016<br><br>UK(24) | Not reported | 5,318 | Mean 41<br>Range not given | Not specified | Swab | Not reported | Cobas 4800 | PreservCyt |
| Leeman 2017<br><br>The Netherlands(28) | Not reported | 91 | Not specified | Not specified | Brush | Clinical setting | SPF10-DEIA-LIPA25 & GP5+/6+-EIA-LMNX | Dry up to 3 months, then placed in vial with PreservCyt for shipment |
| Asciutto 2018<br><br>Sweden(19) | Not reported | 176 | Mean 34<br>Range not given | Not specified | Swab | Clinical setting | APTIMA | APTIMA vaginal specimen collection kit |
| Leinonen 2018<br><br>Norway(30) | Not reported | 240 | Mean 38<br>Range not given | Not specified | Brush | Home | Anyplex II HPV28; cobas 4800, Xpert HPV | Dry transport of self-collection devices to lab |

**Supplemental Table 4: Characteristics of Included Studies on Concordance between HPV-DNA Testing in Self and Health Professional Collected Samples**

| Author, Year, Country | Population | Sample Size | Age (years) | Ethnicity | Device | Setting | hrHPV Assay | Storage medium |
|---|---|---|---|---|---|---|---|---|
| Morrison 1992 USA(55)* | Colposcopy referral | 25 | Not specified | Not specified | Lavage | Clinical setting | PCR | Ethanol carbowax |
| Hillemann 1999 Germany(56)*^1 | Colposcopy referral | 247 | Not specified | Not specified | Lavage | Clinical setting | PCR | Ethanol carbowax |
| Nobbenhuis 2002 The Netherlands(16)*^1 | Colposcopy referral | 71 | Mean 35 Range not given | Not specified | Brush | Clinical setting | PCR | PBS |
| Brink 2006 The Netherlands(17)*^1 | Colposcopy referral | 96 | Median 35 Range 18-59 | Not specified | Brush | Clinical setting | PCR | STM |
| Seo 2006 | Colposcopy referral | 118 | Mean 46.2 | Not specified | Swab | Clinical setting | hrHPV DNA Chip | Not specified |

| **Author, Year, Country** | **Population** | **Sample Size** | **Age (years)** | **Ethnicity** | **Device** | **Setting** | **hrHPV Assay** | **Storage medium** |
|---|---|---|---|---|---|---|---|---|
| South Korea(57)*^1 | | | | | | | | |
| van Baars 2012<br><br>The Netherlands(21)*^1 | Colposcopy referral | 134 | Mean 40<br><br>Range not given | Not specified | Brush | Clinical setting | PCR | FTA cartridge |
| Darlin 2013<br><br>Sweden(58)*^1 | Colposcopy referral | 108 | Mean 34<br><br>Range not given | Not specified | Brush | Clinical setting | PCR | PreservCyt |
| Jentschke 2013a<br><br>Germany (22) | Colposcopy referral | 72 | Mean 37<br><br>Range not given | Not specified | Lavage | Clinical setting | HC2 P16: p16INK4a ELISA | Buffered saline |
| Jentschke 2013b<br><br>Germany(23) | Colposcopy referral | 49 | Mean 36<br><br>Range not given | Not specified | Lavage | Clinical setting | HC2 P16: p16INK4a ELISA | Buffered saline |

| **Author, Year, Country** | **Population** | **Sample Size** | **Age (years)** | **Ethnicity** | **Device** | **Setting** | **hrHPV Assay** | **Storage medium** |
|---|---|---|---|---|---|---|---|---|
| Chernesky 2014<br><br>Canada(59)*^[1] | Colposcopy referral | 580 | Mean 39<br>Range not given | Not specified | Brush | Clinical setting | APTIMA HPV | APTIMA SCT |
| Jentschke 2016<br><br>Germany(25)*^[1] | Colposcopy referral | 136 | Mean 36<br>Range not given | Not specified | Brush | Clinical setting | Abbott RealTime and hrHPV PCR | Dry, then transferred to PreservCyt |
| Aiko 2017<br><br>Japan(26)*^[1] | Colposcopy referral | 136 | Not specified | Not specified | Brush | Clinical setting | HC2 | Not reported |
| Asciutto 2017<br><br>Sweden(27)*^[1] | Colposcopy referral | 218 | Mean 35<br>Range not given | Not specified | Swab | Clinical setting | Cobas 4800 | Cobas PCR Female Swab Sample Kit |
| Catarino 2017<br><br>Switzerland(29)*^[1] | Colposcopy referral | 150 | Mean 32<br>Range not given | Not specified | Swab | Clinical setting | Xpert HPV; part of clin sample also cobas 4800. | Dry samples |
| Leinonen 2018 | Not reported | 240 | Mean 38 | Not specified | Brush | Home | Anyplex II HPV28; cobas | Dry transport of self-collection devices to lab |

| Author, Year, Country | Population | Sample Size | Age (years) | Ethnicity | Device | Setting | hrHPV Assay | Storage medium |
|---|---|---|---|---|---|---|---|---|
| Norway(30)*^1 | | | Range not given | | | | 4800, Xpert HPV | |
| Igidbashian 2014<br>Italy (31) | Not reported | 700 | Mean: 44.3<br>Range not given | Not specified | Not reported | Clinical setting | Hybrid Capture (HC) | Not reported |
| Des Marais 2018<br>USA (60)*^1 | Low income | 193 | Mean 45<br>Range 30–63 | Black (25.7%), White (44.5%), Hispanic (25.7%), Others (4.2%) | Brush | Home | Aptima HPV assay (Hologic, Inc.) | Aptima sample transport media |
| Svensson 2018<br>Sweden (33)*^1 | | 63 | Mean 42<br>Range 24-64 | Not specified | Qvintip | Clinical setting | PCR | Not reported |
| El-Zein 2018<br>Canada (34)^1 | Women referred for colposcopy | 1076 | Mean not Reported<br>Range 21-74 | Not specified | Swab | Clinical setting | PCR | PreservCyt |
| Onuma 2020<br>Japan(37)*^1 | (1) Outpatients with abnormal cytology and requiring colposcopy and biopsy and (2) NILM/HPV-positive | 100 | Mean 41.8<br>Range not given | Not specified | Brush | Clinical setting | PCR | ThinPrep vials |

| Author, Year, Country | Population | Sample Size | Age (years) | Ethnicity | Device | Setting | hrHPV Assay | Storage medium |
|---|---|---|---|---|---|---|---|---|
| | patients in the Fukui Cervical Cancer Study | | | | | | | |
| Woong Cho 2020<br><br>South Korea(39)* | Women referred to colposcopy for abnormal cytology | 314 | 40±15.4 years (Reported this a median age) | | Swab | Clinical Setting | PCR | PreservCyt Solution (ThinPrep) |
| Rohner 2020b<br><br>USA(41)*^1 | Women who were attending colposcopy clinics | 307 | Median 36<br><br>Range not given | Non-Hispanic white 38%; Hispanic white 29%; Non-Hispanic 26%; other (7%) | Not reported | Not reported | PCR (Urine sample) | Becton Dickinson (BD) molecular tube containing 0.2 ml of a proprietary preservative |
| Satake 2020<br><br>Japan(61)* | No details provided | 300 | Mean not reported<br><br>Range 20-59 | Not specified | Home Smear Set (ISK Co., Ltd., Tokyo, Japan) | Clinical setting | PCR | Cell fixation container (principal component is ethanol) |

| **Author, Year, Country** | **Population** | **Sample Size** | **Age (years)** | **Ethnicity** | **Device** | **Setting** | **hrHPV Assay** | **Storage medium** |
|---|---|---|---|---|---|---|---|---|
| Saville 2020 Australia(62)*^1 | Referral for colposcopy | 292-296 | Not reported | Not specified | Swab | Clinical setting | Cobas 4800; Cobas; Onclarity; GeneXpert; Anyplex II; Abbott | Not reported |
| Tranberg 2020 Denmark(63)*^1 | Women diagnosed with ASC-US. | 150 | Median 45 Range not given | Not specified | Not specified | Home | GENOMICA CLART® Cobas | Transportation tube with preservative media (Genelock, ASSAY ASSURE, Sierra Molecular, CA |
| Ertik 2021 Germany(64) | Patients referred to colposcopy clinics with abnormal results | 65 | Mean 36 Range, 24–76 | Not specified | Swab, Brush | Home | PCR | ThinPrep PreservCyt |
| Hong Kim 2021 South Korea(65) | Women who had abnormal cervical smears or who were HPV-positive | 151 | Median 50 Range 21–65 | Not specified | G+Kit®; DocTool | Clinical setting | PCR | Not reported |
| Klischke 2021 Germany(42) | Patients from the colposcopy clinic | 70 | Mean 37 Range not given | Not specified | Brush | Clinical setting | PCR | ThinPrep PreservCyt Solution |

| **Author, Year, Country** | **Population** | **Sample Size** | **Age (years)** | **Ethnicity** | **Device** | **Setting** | **hrHPV Assay** | **Storage medium** |
|---|---|---|---|---|---|---|---|---|
| Rohner 2021<br>USA(66)*^1 | Women attending colposcopy clinics with i) abnormal cytology results, ii) infection with HPV-16 or 18, iii) persistent infection with other hr-HPV genotypes, or iv) treatment for CIN2+ | 314 | Median 36<br>Range not given | Non-Hispanic white 38%; Hispanic 29%; non-Hispanic black 26% and others 6% | Brush | Not reported | PCR | ThinPrep |
| Avian 2022<br>Italy(44) | | 889 | Not specified | Not specified | Swab | Clinical setting | PCR | ThinPrep |
| Giubbi 2022<br>Italy(67) | Women, referred to colposcopy | 30 | Mean 36.5<br>Range not given | Not specified | Swab | Clinical setting | PCR (Anyplex™II HPV28 (Seegene); HPV28 (Seegene)); Papilloplex® High Risk HPV; (GeneFirst); HPV OncoPredict (Hiantis) | ThinPrep®PreservCyt®; eNat® |

| Author, Year, Country | Population | Sample Size | Age (years) | Ethnicity | Device | Setting | hrHPV Assay | Storage medium |
|---|---|---|---|---|---|---|---|---|
| Martinelli 2022<br>Italy(68)*^2 | Women referred to colposcopy | 64 | Mean 38.4<br>Range not given | Not specified | Swab<br>Colli-pee®- for first-void urine (FVU) sample | Not specified | BD Onclarity™ HPV Assay | PreservCyt<br>Preservative urine conservation medium (UCM) |
| Naseri 2022<br>USA(69)*^2 | Women with and without a history of high-risk HPV infection and with regular menses | 106 | Mean<br>31.0<br>Range not given | Asian 35.8%; Black 1.9%; Native Hawaiian/Other Pacific Islander 1.9%; White 48.1%, others 11.3% | Swab<br>Q-Pad (QvinTM, Menlo Park, CA) | Clinical setting<br>Home | Roche Cobas 4,800 | Cobas media solution.<br>Dry samples |
| Ngu 2022<br>Hong Kong(70)*^1 | History of sexual activity and underserved population | 121 | Mean not reported<br>Range 30-65 | Not specified | Swab | Not reported | PCR | PreservCyt media |
| Terada 2022<br>Japan(71) | Women attending hospital for abnormal cervical cytology | 300 | Mean not reported | Not specified | Brush | Not reported | PCR | PreservCyt |

| Author, Year, Country | Population | Sample Size | Age (years) | Ethnicity | Device | Setting | hrHPV Assay | Storage medium |
|---|---|---|---|---|---|---|---|---|
| | | | Range 21-50 | | Colli-pee®- for urine (FVU) sample | | | |
| Stanczuk 2022<br>UK(46) | Women eligible for cervical screening | 4617 | Mean 41.3<br>Range not given | Not specified | Not specified | | Cobas 4800 PCR-based DNA test | ThinPrep (PreservCyt Solution, Holgic UK) |
| Gibert 2023<br>Spain(72)^1 | Women recruited from a colposcopy clinic | 120 | Median 46<br>Range 40–51 | Spain 62.5%; Central and South America 21.7%; European and United Kingdom 7.5%, Others (8.3%) | Swab, Iune HPV sterile test cannula, brush, Mia by XytoTest | Clinical setting | PCR | PreservCyt, reTect TM Preservation and Transport Media |
| Martinelli 2024<br>Italy(50)^1 | Women who were referred to colposcopy | 286 | Median 40<br>Range not given | Not specified | Swab | Clinical sampling | Ist sample on VIPER; Second vaginal sample with VIPER; Second vaginal sample with COR | Dry samples |

* and ^ indicate studies that were included in the meta-analysis for overall agreement and kappa respectively. ^[1] and ^[2] indicate that the variance for kappa was directly taken from the study or calculated from other data respectively.

Coloured red: studies from top-up search

**Supplemental Table 5: Characteristics of Studies Included Studies for Uptake of HPV DNA Self Sampling that had no outcome in the reference review**

| Author, Year, Country | Population | Sample Size | Age (years) | Invitation Strategy | Reminder | Time from Invitation to collected sample | Self-sampling device | Per protocol (PP) or Intention to Treat (ITT) Analysis | Outcomes Reported |
|---|---|---|---|---|---|---|---|---|---|
| Veerus 2021<br><br>Estonia(73) | Never screened; Under screened | Intervention Mail-to-all: 4000 Opt-in: 8000<br><br>Comparator Not started | Range 37-62 | Mail-to-all; Opt-in | No | Not documented | Qvintip and Evalyn brush | Not reported | Not reported |

### Supplemental Table 6: Characteristics of Included Studies for Uptake Question

| **Author, Year, Country** | **Population** | **Sample Size** | **Age (years)** | **Invitation Strategy** | **Reminder** | **Time from Invitation to collected sample** | **Self-sampling device** | **Per protocol (PP) or Intention to Treat (ITT) Analysis** | **Outcomes Reported** |
|---|---|---|---|---|---|---|---|---|---|
| Bais<br>2007<br>New Zealand(74) | Under screened | Intervention 2,352<br>Comparator 272 | Range 30-50 | Mail to all | No | 6 months | Brush | PP &ITT | Response rates; adherence to follow-up; insufficient sample; CIN+ 2 detection |
| Gok<br>2010<br>The Netherlands (75) | Under screened | Intervention<br>26,886<br>Comparator 277 | Range 30-60 | Mail-to-all | | 12months | Lavage | PP &ITT | Response rates; adherence to follow-up; insufficient sample; CIN+ 2 detection |
| Giorgi-Rossi<br>2011<br>Italy(76) | | Intervention<br>Mail-to-all: 616; Opt-in: 622<br>Comparator | Range 35-65 | Mail-to-all; Opt-in | No | 3 months | Brush | PP &ITT | Response rates; adherence to follow-up; insufficient sample; CIN+ 2 detection |

| Author, Year, Country | Population | Sample Size | Age (years) | Invitation Strategy | Reminder | Time from Invitation to collected sample | Self-sampling device | Per protocol (PP) or Intention to Treat (ITT) Analysis | Outcomes Reported |
|---|---|---|---|---|---|---|---|---|---|
| | | Mail-to-all: 619; Opt-in: 616 | | | | | | | |
| Piana, 2011 France(77) | Under screened | Intervention 4,400 Comparator 4,934 | Range 35-69 | Mail-to-all | No | Not documented | Not documented | PP &ITT | Response rates; adherence to follow-up; insufficient sample; CIN+ 2 detection |
| Szarewski 2011 UK(78) | Under screened | 1,500 in both intervention and comparator | Range 25-64 | Mail-to-all | No | 6 months | Swab | PP &ITT | Response rates; adherence to follow-up; insufficient sample; CIN+ 2 detection |
| Virtanen 2011 Finland(79) | Under screened | Intervention 2,397 Comparator | Range 30-60 | Mail-to-all | No | Not documented | Lavage | PP &ITT | Response rates; adherence to follow-up; insufficient sample; CIN+ 2 detection |

| **Author, Year, Country** | **Population** | **Sample Size** | **Age (years)** | **Invitation Strategy** | **Reminder** | **Time from Invitation to collected sample** | **Self-sampling device** | **Per protocol (PP) or Intention to Treat (ITT) Analysis** | **Outcomes Reported** |
|---|---|---|---|---|---|---|---|---|---|
| | | 6,302 | | | | | | | |
| Wikstrom<br>2011<br>Sweden(80) | Under screened | Intervention<br>2,000<br>Comparator<br>2,060 | Range 39-60 | Mail-to-all | Yes | 12 months | Swab | PP &ITT | Response rates; adherence to follow-up; CIN+ 2 detection |
| Gok<br>2012<br>The Netherlands (81) | Under screened | Intervention<br>25,561<br>Comparator<br>261 | Range 30-60 | Mail-to-all | No | 12 months | Brush | PP &ITT | Response rates; adherence to follow-up; insufficient sample; CIN+ 2 detection |
| Darlin<br>2013<br>Sweden(82) | Under screened | Intervention<br>1000<br>Comparator | Range 32-65 | Mail-to-all | Yes | Not documented | Not documented | PP &ITT | Response rates; adherence to follow-up; insufficient sample; CIN+ 2 detection |

| Author, Year, Country | Population | Sample Size | Age (years) | Invitation Strategy | Reminder | Time from Invitation to collected sample | Self-sampling device | Per protocol (PP) or Intention to Treat (ITT) Analysis | Outcomes Reported |
|---|---|---|---|---|---|---|---|---|---|
| | | 500 | | | | | | | |
| Sancho-Garnier<br>2013(83)<br>France(83) | Under screened | Intervention<br>8,829<br>Comparator<br>9,901 | Range 35-69 | Mail-to-all | No | Not documented | Swab | PP &ITT | Response rates; adherence to follow-up; insufficient sample; CIN+ 2 detection |
| Broberg<br>2014<br>Sweden(84) | Never screened; Under screened | Intervention<br>800<br>Comparator<br>4000 | Range 30-62 | Opt-in | Yes | Not documented | Swab | PP &ITT | Response rates; adherence to follow-up; CIN+ 2 detection |
| Haguenoer<br>2014<br>France(85) | Under screened | Intervention<br>1,999<br>Comparator<br>Cytology 2,000 | Range 30-65 | Mail-to-all | No | 9m; 12m | Swab | PP &ITT | Response rates; adherence to follow-up; insufficient sample; CIN+ 2 detection |

| **Author, Year, Country** | **Population** | **Sample Size** | **Age (years)** | **Invitation Strategy** | **Reminder** | **Time from Invitation to collected sample** | **Self-sampling device** | **Per protocol (PP) or Intention to Treat (ITT) Analysis** | **Outcomes Reported** |
|---|---|---|---|---|---|---|---|---|---|
| | | No intervention 1,999 | | | | | | | |
| Cadman<br>2015<br>UK(86) | Under screened | 3000 in both arm | Range 25-65 | Mail-to-all | No | 3 months | Swab | PP &ITT | Response rates; adherence to follow-up; insufficient sample; CIN+ 2 detection |
| Giorgi-Rossi<br>2015<br>Italy(87) | Under screened | Intervention<br>Mail-to-all: 4,516; Opt-in: 4,513<br>Comparator<br>Mail-to-all: 1,998; Opt-in: 3,014 | Range 30-64 | Mail-to-all; Opt-in | No | 3 months | Lavage | PP &ITT | Response rates; adherence to follow-up; insufficient sample; CIN+ 2 detection |
| Enerly<br>2016 | Under screened | Intervention<br>800 | Range 26-69 | Mail-to-all | No | Not documented | Lavage (Delphi screener) / | PP &ITT | Response rates; adherence to follow-up; |

| Author, Year, Country | Population | Sample Size | Age (years) | Invitation Strategy | Reminder | Time from Invitation to collected sample | Self-sampling device | Per protocol (PP) or Intention to Treat (ITT) Analysis | Outcomes Reported |
|---|---|---|---|---|---|---|---|---|---|
| Norway(88) | | Comparator<br>2,593 | | | | | Evalyn brush (randomized) | | insufficient sample |
| Sultana<br>2016<br>Australia(89) | Never screened; Under screened | Intervention<br>14,153 (7,075 un-screened; 7,078 under-screened)<br>Comparator<br>2,025 (1,014 un-screened; 1,011 under- screened) | Range 30-69 | Mail-to-all | No | 6 months | Swab | PP &ITT | Response rates; adherence to follow-up; insufficient sample; CIN+ 2 detection |
| Kitchener<br>2017<br>UK(90) | Under screened | Intervention<br>Mail-to-all: 1,141 (32 GPs); Opt-in: 1,290 (66 GPs) | Mean 20 (Grampian)<br>Mean 25 (Manchester)) | Mail-to-all; Opt-in | No | 3m, 6m, 12m, 18m | Lavage (Delphi Screener)/ Evalyn Brush | PP &ITT | Response rates. |

| Author, Year, Country | Population | Sample Size | Age (years) | Invitation Strategy | Reminder | Time from Invitation to collected sample | Self-sampling device | Per protocol (PP) or Intention to Treat (ITT) Analysis | Outcomes Reported |
|---|---|---|---|---|---|---|---|---|---|
| | | Comparator<br>3,782 (101 GPs) | | | | | | | |
| Kellen<br>2018<br>Belgium(91) | Under screened | Intervention<br>Mail-to-all: 9,118; Opt-in: 9,098.<br>Comparator<br>Reminder letter: 8,830; No reminder: 8,849 | Range 30-64 | Mail-to-all; Opt-in | Yes | 12m | Qvintip | PP &ITT | Response rates. |
| Tranberg<br>2018<br>Denmark(92) | Never screened; Under screened | Intervention<br>Mail-to-all: 3,265; Opt-in: 3,264.<br>Comparator<br>3,262 | Range 30-64 | Mail-to-all; Opt-in | Yes | 6 months | Brush | PP &ITT | Response rates; adherence to follow; CIN+ detection |

| **Author, Year, Country** | **Population** | **Sample Size** | **Age (years)** | **Invitation Strategy** | **Reminder** | **Time from Invitation to collected sample** | **Self-sampling device** | **Per protocol (PP) or Intention to Treat (ITT) Analysis** | **Outcomes Reported** |
|---|---|---|---|---|---|---|---|---|---|
| Ivanus 2018 Slovenia(93) | Under screened | Intervention Mail-to-all: 9,556; Opt-in: 14,400 Comparator 2600 | Range 34-64 | Mail-to-all; Opt-in | No | 12 months | Mail-to-all: Qvintip (Swab), HerSwab (Swab) and Delphi Screener (Lavage). Opt-in: Qvintip | PP &ITT | Response rates; adherence to follow-up |
| Elfström 2019 Sweden(94) | Under screened | Intervention Mail-to-all: 2,000; Opt-in: 2,000 Comparator 2000 | Range 33 - 60 | Mail-to-all; Opt-in | No | 3 months | Swab | PP &ITT | Response rates; CIN+ detection |
| Jalili 2019 | Under screened | Intervention 529 Comparator | Range 30 - 65 | Mail-to-all | Yes | 6 months | Swab | PP &ITT | Response rates |

| Author, Year, Country | Population | Sample Size | Age (years) | Invitation Strategy | Reminder | Time from Invitation to collected sample | Self-sampling device | Per protocol (PP) or Intention to Treat (ITT) Analysis | Outcomes Reported |
|---|---|---|---|---|---|---|---|---|---|
| Canada(95) | | 523 | | | | | | | |
| Winer<br>2019<br>USA(96) | Under screened | Intervention<br>9,960<br>Comparator<br>9,891 | 30 - 64 | Mail-to-all | No | 6months | Not documented | PP &ITT | Response rates; adherence to follow-up; insufficient sample; CIN+ 2 detection |
| Lilliecreutz<br>2020<br>Sweden(97) | Under screened | Intervention<br>3,068<br>Comparator<br>3,538 | Range 30 - 64 | Mail-to-all | Yes | 6 months | Swab | PP &ITT | Response rates; adherence to follow-up; CIN+ detection |
| Brewer<br>2021 | Never screened; | Intervention | Range 30-69 | Mail-to-all; Opt-in, and | Yes | 3 months | Swab | PP &ITT | Response rates; follow up; |

| Author, Year, Country | Population | Sample Size | Age (years) | Invitation Strategy | Reminder | Time from Invitation to collected sample | Self-sampling device | Per protocol (PP) or Intention to Treat (ITT) Analysis | Outcomes Reported |
|---|---|---|---|---|---|---|---|---|---|
| New Zealand(98) | Under screened | Mail-to-all: 1467: Opt-in: 1574<br><br>Comparator<br><br>512 | | Opportuni stic | | | | | insufficient sample |
| Virtanen<br><br>2014<br><br>Finland(79) | Under screened, never screened | Intervention<br><br>4536<br><br>Comparator<br><br>Not reported | Range 25-67 | Mail-to-all | Not reported | Not documented | Lavage | Not reported | Response rates; adherence to follow-up; CIN2+ |
| Lam<br><br>2017<br><br>Denmark(99) | Under-screened, never screened | Intervention 23,632 | Range 27 - 65 | Opt-in | Yes | 8 weeks | Brush | PP and ITT | Response rates |
| Gunvor Aasbø | Never screened; | 2000 in both arms | Mean 54.3 | Mail-to-all; Opt-in | Yes | Not documented | Brush | PP &TT | Response rates; adherence to |

| **Author, Year, Country** | **Population** | **Sample Size** | **Age (years)** | **Invitation Strategy** | **Reminder** | **Time from Invitation to collected sample** | **Self-sampling device** | **Per protocol (PP) or Intention to Treat (ITT) Analysis** | **Outcomes Reported** |
|---|---|---|---|---|---|---|---|---|---|
| 2022<br><br>Norway(100) | Under screened | | | | | | | | follow-up; CIN2+ detection |
| Fujita<br><br>2022<br><br>Japan(101) | Never screened; Under screened | Intervention<br>7,340<br><br>Comparator<br>7,782 | Range 30-59 | Opt-in | Yes | Not documented | Brush | Not reported | Response rates; insufficient sample |
| Ejegod<br><br>2022<br><br>Denmark(102) | Never screened; Under screened | Intervention<br>57,717<br><br>Comparator<br>Not reported | Range 27-65 | Opt-in | Yes | Not documented | Brush | PP & ITT | Response rates; adherence to follow-up |
| Sultana<br><br>2022 | Never screened; Under screened | Intervention<br>12,572 | Range 30-69 | Mail-to-all | No | 2 years | Swab | Not reported | Response rates; adherence to follow-up; insufficient |

| Author, Year, Country | Population | Sample Size | Age (years) | Invitation Strategy | Reminder | Time from Invitation to collected sample | Self-sampling device | Per protocol (PP) or Intention to Treat (ITT) Analysis | Outcomes Reported |
|---|---|---|---|---|---|---|---|---|---|
| Australia(103) | | Comparator<br>Not reported | | | | | | | sample; CIN+ 2 detection |
| Winer<br>2022<br>USA(104) | Never screened; Under screened.<br>White 71.6%, did not specify others' percentage | Intervention<br>9843<br>Comparator<br>9891 | Mean 50.1 | Mail-to-all | Not reported | Enrolled for 3 years and 5 months or more, and with no Papanicolaou test within 3 years and 5 months | Not reported | ITT | Response rates |
| Auvinen<br>2022<br>Finland(105) | | Intervention<br>5350<br>Comparator<br>Not reported | Range 25-69 | Opt-in. | Not reported | No documented | Aptima Multitest sampling kit | Not documented | Response rates; adherence to follow-up; insufficient sample; CIN+ 2 detection |
| Nishimura 2023 | Never screened; | Intervention<br>7,653 | Range 20-50 | Opt in | Not reported | Not documented | Brush | ITT | Response rates' adherence to |

| Author, Year, Country | Population | Sample Size | Age (years) | Invitation Strategy | Reminder | Time from Invitation to collected sample | Self-sampling device | Per protocol (PP) or Intention to Treat (ITT) Analysis | Outcomes Reported |
|---|---|---|---|---|---|---|---|---|---|
| Japan(106) | Under screened | No Comparator | | | | | | | follow-up; CIN2+ |
| Winer 2023 USA(107) | Never screened; Under screened; Routinely screened Due for screening: White 73.4%; Asian 12.4%; Black or African American 4.9%; others 9.3% Overdue: White 73.6%; Asian | Intervention Due for screening 12,928; Overdue for screening 8279; Unknown screening history 9942 Comparator 12,142 | Mean 45.9 | Opt in; Mail-to-all | Yes | Due for screening ≤3 months; Overdue for screening (co-testing >5.25years ago, Papanicolaou testing alone >3.25 years ago, or no Papanicolaou testing with continuous enrolment ≥3.25 years, unknown enrolment ≥6 months and <3.25 years, | Swab | ITT | Response rates; adherence to follow-up |

| **Author, Year, Country** | **Population** | **Sample Size** | **Age (years)** | **Invitation Strategy** | **Reminder** | **Time from Invitation to collected sample** | **Self-sampling device** | **Per protocol (PP) or Intention to Treat (ITT) Analysis** | **Outcomes Reported** |
|---|---|---|---|---|---|---|---|---|---|
| | 11.5%; Black or African American 5.2%; others 9.7% | | | | | no recorded screening) | | | |
| Taro2024<br><br>Japan(108) | Never screened; Under screened | Intervention<br>3489<br><br>Comparator<br>Not reported | 30-39 | Opt-in | Not reported | Not documented | Brush | PP & ITT | Response rates,<br>Adherence to follow-up; CIN2+ |
| Ngo2024<br><br>Czech Republic(109) | Never screened; Under screened | Intervention<br>800<br><br>Comparator<br>764 | Range 50–65 | Mail-to-all | Yes | Not documented | Brush | PP & ITT | Response rates' insufficient sample; dherence to follow-up |

| Author, Year, Country | Population | Sample Size | Age (years) | Invitation Strategy | Reminder | Time from Invitation to collected sample | Self-sampling device | Per protocol (PP) or Intention to Treat (ITT) Analysis | Outcomes Reported |
|---|---|---|---|---|---|---|---|---|---|

**NB**: Response rates: if the study reported any of the following absolute response rate, relative response rate, response difference. Adherence to follow-up: if the study reported on adherence to follow-up of individuals who receive positive screening results. Insufficient sample: proportion of individuals with unsatisfactory test results i.e HPV status could not be determined.

Colored red: studies from top-up search.

**Supplemental Table 7: Characteristics of Included Studies for Acceptability of HPV Self-sampling Screening Included in reference review without outcomes**

| Author, Year, Country | Population | Sample size | Age (years) | Invitation Strategy | Self-sampling device used | Outcomes | |
|---|---|---|---|---|---|---|---|
| | | | | | | Acceptability | Individual characteristics of acceptability |
| Harper 2002<br><br>USA(110) | | 67 | Mean 37.7 | | Dacron Swab and Tampon | | |
| Jones 2012<br><br>USA(111) | | 197 | Median 45 | | Lavage | | |
| Litton 2013<br><br>USA(112) | | 516 | ≥30 | | Not reported | | |
| Chen 2014<br><br>Taiwan(113) | | 297 | Range 18-65 | | Unable to determine | | |
| Haguenoer 2014<br><br>France(85) | | 722 | Range 20-65 | | Swab | | |

**Supplemental Table 8 Characteristics of Included Studies for Acceptability of HPV Self-sampling Screening Strategies**

| Author, Year, Country | Population | Sample size | Age (years) | Invitation Strategy | Self-sampling device used | Outcomes | |
|---|---|---|---|---|---|---|---|
| | | | | | | Acceptability | Individual characteristics of acceptability |
| Dannecker 2004<br>Germany(114) | | 333 | Mean 45 | | Brush | Overall acceptability; preference | |
| Kahn 2005<br>USA(115) | | 120 | Mean 17.8<br>Range 14-21 | | Swab | Preference | |
| Anhang 2005<br>USA(116) | | 172 | 25%: 25-35; 10%: >55 | Not specified | Swab | Preferences | |
| Waller 2006<br>UK(117) | | 902 | Mean 34.2 | | Swab | Preference | |
| Wikstrom 2007<br>Sweden(118) | | 94 | Range 35-55 | | Qvintip | Preference | |
| Barbee 2010 | | 245 | 6%:18-25; 94%: ≥25 | | Tampon | Preference | |

| Author, Year, Country | Population | Sample size | Age (years) | Invitation Strategy | Self-sampling device used | Outcomes | |
|---|---|---|---|---|---|---|---|
| | | | | | | Acceptability | Individual characteristics of acceptability |
| USA(119) | | | | | | | |
| Cerigo 2011<br>Canada(120) | | 92 | Mean 33.2<br>Range 18-69 | | Swab | Preference | |
| Delere 2011<br>Germany (121) | | 156 | Range 20-30 | | Lavage | | |
| Igidbashian 2011<br>Italy(122) | | 194 | Mean 39.6<br>Range 19-72 | | Brush and Delphi screener (Lavage) | Overall acceptability; preference | |
| Rossi 2011<br>Italy(76) | | 147 | Range 25-64 | | Not reported | Preference | |
| Ortiz 2012<br>Puerto Rico(123) | | 100 | Mean 26.4<br>Range 18-34 | | Dacron Swab, CytoBrush | Preference | |
| Van Baars 2012 | | 127 | Median 40 | | Brush | Overall acceptability; preference | |

| Author, Year, Country | Population | Sample size | Age (years) | Invitation Strategy | Self-sampling device used | Outcomes | |
|---|---|---|---|---|---|---|---|
| | | | | | | Acceptability | Individual characteristics of acceptability |
| The Netherlands(21) | | | | | | | |
| Castell 2014<br><br>Germany(124) | | 108 | Range 20-69 | | Lavage | Overall acceptability; preference | |
| Catarino Jr 2014<br><br>Switzerland(125) | | 158 | Mean 43.6 | | Swab | Overall acceptability; preference | |
| Montealegre 2014<br><br>USA(126) | | 100 | Median 38 | | Cytology Broom | Acceptability | |
| Nelson 2014<br><br>USA(127) | | 67 | Median 24<br><br>Range 21-30 | | Swab | Preference | |
| Virtanen 2014<br><br>Finland(128) | Finish 93%; Swedish 2.2%; Other 4.8% | 909 | Range 30-64 | | Lavage | | Procedural and psychosocial |

| Author, Year, Country | Population | Sample size | Age (years) | Invitation Strategy | Self-sampling device used | Outcomes | |
|---|---|---|---|---|---|---|---|
| | | | | | | Acceptability | Individual characteristics of acceptability |
| Vanderpool 2014 USA (Appalachian)(129) | Low income Caucasian (100%) | 31 | Mean 38.5 | | Brush | Overall acceptability | |
| Galbraith 2014 USA(130) | Low-income status women: Non-Hispanic Black (55%), White (33%), Other (13%) | 199 | Range 30-65 | | Brush | Overall acceptability; preference | Procedural |
| Bosgraaf 2014 The Netherlands(131) | | 9484 | Range 29-63 | | Lavage and brush | Preference | Logistic and psychosocial |
| Catarino 2015 Switzerland(132) | European (39.8%), Swiss (17.7%), Asian (7.0%), African (9.5%), Latin American (36.7%), Others (7.0%) | 158 | Mean 43.6 | | Swab | Overall acceptability; preference | Procedural |
| Chou 2015 Taiwan(133) | | 282 | Mean 48.1 | Mail-to-all | Brush | Overall acceptability | Procedural |
| Crosby 2015 | Rural, economically disadvantaged area: White (93.8%), | 400 | Mean 40.2 | Community outreach and mobilization | Swab | Preference | procedural |

| Author, Year, Country | Population | Sample size | Age (years) | Invitation Strategy | Self-sampling device used | Outcomes | |
|---|---|---|---|---|---|---|---|
| | | | | | | Acceptability | Individual characteristics of acceptability |
| USA (rural Appalachian)(134) | Black (2.8%), and others (3.4) | | | | | | |
| Sultana 2015<br>Australia(135) | | 746 | 30-69 (inclusion criteria) | | Swab | Preference | Logistic, procedural and psychosocial |
| Crosby 2016<br>USA(136) | A highly impoverished and geographically isolated population of medically underserved Black women residing in the Mississippi Delta | 88 | Mean 46.5 | Community outreach and mobilization | Swab | Preference | Procedural |
| Ilangovan 2016<br>USA(137) | Women in Safety Net institutions: Latinas (74.4%), Haitian (25.6%) | 180 (those who completed the questionnaire for self-sampling were 121) | Mean 52 | Offered in the healthcare setting | Preventive Oncology International/ National Institute of Health self-sampler | Preference | Logistic, procedural, and psychosocial |
| Racey 2016<br>Canada(138) | | 70 | Mean 53.6 Range 51.2-56.0 | | Swab | Overall acceptability; preference | |

| Author, Year, Country | Population | Sample size | Age (years) | Invitation Strategy | Self-sampling device used | Outcomes | |
|---|---|---|---|---|---|---|---|
| | | | | | | Acceptability | Individual characteristics of acceptability |
| Levinson 2016<br>USA(139) | White (59%), Black (41%) | 35 | Median 38 | | | Preference | |
| Anderson 2017<br>USA(140) | Low income:<br>Black (55%), White (35%), Other (10%) | 227 | Median 44<br>Range 30-64 | | Brush | Overall acceptability; preference | Logistic and procedural |
| Karjalainen 2016<br>Finland(141) | | 67 (39 lavage, 28 Brush) | | | Lavage and Brush | | Logistic, procedural, and psychosocial |
| Kilfoyle 2018<br>USA(142) | Low-income women: White (35%), Black (56%), and others (9%) | 221 (the acceptance was reported for 100) | Median 44<br>Range 30–64 | | | Overall acceptability; preference | Procedural, and psychosocial |
| Des Marais 2018<br>USA(60) | Low-income women: White (45%), Black (26%), Hispanic (26%), Other races (4%) | 193 | Median age 45<br>Range 30-63 | | Brush | Overall acceptability; preference | Procedural |
| Molokwu 2018<br>USA(143) | | 202 | Mean 46.4 | Community outreach and mobilization | | Preference | |

| Author, Year, Country | Population | Sample size | Age (years) | Invitation Strategy | Self-sampling device used | Outcomes | |
|---|---|---|---|---|---|---|---|
| | | | | | | Acceptability | Individual characteristics of acceptability |
| Smith 2018<br>USA(100) | Low income | 227 | Median 42<br>Range 30-65 | | Brush | Overall acceptability | |
| Brewer 2019<br>New Zealand(144) | Pacific (55.4), Maori (21.4), Asian (16.1), other (7.1) | 56 (herSwab N=51, Delphi Screener 8, Cobas CT/NG Swab 7) | Median 39.5<br>Range 20-61 | Opt-in; Mail-to-all | Swabs and Delphi Screener (Rovers Medical Devices) | Overall acceptability; preference | Logistic, procedural and psychosocial |
| Adcock 2019<br>New Zealand(145) | Maori (100%) | 397 | ≥25 | | | Overall acceptability; preference | Procedural and psychosocial |
| Reiter 2019<br>USA (Appalachain)(146) | White, non-Hispanic (98%) and others (2%) | 79 | Mean 46.4 | | Brush | Preference | Logistic and psychosocial |
| Datta 2020<br>Canada(147) | Never screeners: Canada (62%), United States/Europe (9%), other countries (28%); Under screeners: Canada (90%), United | Never 53, Under screeners 89 | 21 -65 (Inclusion criteria) | | | Overall acceptability | |

| Author, Year, Country | Population | Sample size | Age (years) | Invitation Strategy | Self-sampling device used | Outcomes | |
|---|---|---|---|---|---|---|---|
| | | | | | | Acceptability | Individual characteristics of acceptability |
| | States/Europe (4%), other countries (6%) | | | | | | |
| Malone 2020<br>USA(148) | White (88.8%), Black/African American (0.9%), Asian/Pacific Islander (5.2%), others (4.3%), and unknown (0.9%) | 120 | Range 30-64 | Mail-to-all | Swab | Preference | Logistic, procedural, and psychosocial |
| Andersson 2021<br>Sweden(149) | | 43 cases, 479 control (controls are not long-term non-attenders hence results are only reported for cases) | Case Mean 44.5 | | Swab | Overall acceptability | Logistic and procedural |
| Bromhead 2021(150) | Māori, Pacific and Asian | 58 | Median 45<br>Range 30-68 | | Swab | Preference | Logistic, procedural and psychosocial |
| Veerus 2021<br>Estonia(151) | | 1857 | Range 37-62 range | Opt-in; Mail-to-all | Qvintip and Evalyn brush | Preference | procedural and psychosocial |

| Author, Year, Country | Population | Sample size | Age (years) | Invitation Strategy | Self-sampling device used | Outcomes | |
|---|---|---|---|---|---|---|---|
| | | | | | | Acceptability | Individual characteristics of acceptability |
| Chaw 2022<br>Brunei(152) | Malay 93.0%, Chinese 4.1%, Other 0.31% | 97 | Median 41 | Offer in the healthcare setting | Brush | Preference | Logistic, procedural and psychosocial |
| Ngu 2022<br>Hong Kong(70) | Chinese (52.3%), Philippine (38.9%), Asian-not specified (4.4%), and unknown (5%) | 321 | Range 30-65 range | Community outreach and mobilization and opt-in | Swab | Overall acceptability; preference | Logistic, procedural, and psychosocial |
| Parker 2022<br>USA(153) | Low income enrolled in the safety net: Mexico (39.5%), United States (20.6%), Central America (20.6%), South America (1.7%), Asia (0.9%), Europe (1.3%) and other (0.9%) | 153 | Mean 47.2 | Mail-to-all | Swab | | Logistic and psychosocial |
| Sherman 2022<br>New Zealand(154) | Maori (28.7%), Pasifika (27.9%), and Asian (43.4%) | 376 | Mean 46.5 | | Swab | Preference | Logistic, procedural and psychosocial |

| Author, Year, Country | Population | Sample size | Age (years) | Invitation Strategy | Self-sampling device used | Outcomes | |
|---|---|---|---|---|---|---|---|
| | | | | | | Acceptability | Individual characteristics of acceptability |
| Zhu 2022<br>Canada(155) | North American Aboriginal (2.5%), Other North American (43.9%), European (31.3%), Asian (17.6%), and other (4.8%) | 524 | Mean 47.9 | | | Overall acceptability | |
| Fujita 2023<br>Japan(156) | | 1,192 | Mean 44.1 | | Brush | | Logistic and psychosocial |

**Supplemental Table 9: Quality of Included Studies: Accuracy of HPV testing in self-collected samples compared with health professional-collected samples**

| Study | Patient selection | | Index test | | Reference standard | | Flow and Timin |
|---|---|---|---|---|---|---|---|
| | Risk of bias | Applicability concern | Risk of bias | Applicability concern | Risk of bias | Applicability concern | Risk of bias |
| Aiko 2017 | Low | High | High | Low | Low | Low | Low |
| Avian 2022 | Low | High | Low | Low | Low | Low | Unclear |
| Cho 2020 | Low | High | Low | Low | Low | Low | High |
| Edblad-Svensson 2018 | Low | High | High | Low | Low | Low | High |
| El-Zein 2018 | Low | High | Low | Low | Low | Low | Unclear |
| El-Zein 2019 | Low | High | Low | Low | Low | Low | Unclear |
| Ertik 2021 | Low | High | Low | Low | Low | Low | Low |
| Igidbashian 2014 | Low | High | Low | Low | Low | Low | High |

| | | | | | | | |
|---|---|---|---|---|---|---|---|
| Klischke 2021 | Unclear | High | Low | Low | Low | Low | High |
| Latsuzbaia 2022a | Low | High | Low | Low | Low | Low | Low |
| Latsuzbaia 2023a | Low | High | Low | Low | Low | Low | Low |
| Latsuzbaia 2022b | Low | High | Low | Low | Low | Low | Low |
| Latsuzbaia 2023b | Low | High | Low | Low | Low | Low | Low |
| Leinonen 2018 | Unclear | High | Low | Low | Low | Low | High |
| Mangold 2019 | Unclear | High | Low | Low | Low | Low | Low |
| Martinelli 2023 | Unclear | High | Unclear | Unclear | Low | Low | Low |
| Martinelli 2024 | Low | High | Low | Low | Low | Low | Low |
| Naseri 2022 | Low | High | Low | High | Low | Low | Low |
| Onuma 2020 | High | High | Low | Low | Low | Low | Low |
| Ornskov 2021 | Low | High | Low | Low | Low | Low | Low |
| Pasquier 2023 | Unclear | High | Low | Low | Low | Low | Low |
| Polman 2019 | Low | Low | Low | Low | Low | Low | High |
| Rohner 2020a | Low | High | Low | Low | Low | Low | High |
| Rohner 2020b | Low | High | Low | Low | Low | Low | High |
| Satake 2020 | Low | High | Low | Low | Low | Low | Low |

### Supplemental Table 10: Quality of Included Studies for Uptake

| RoB Tool | Author, Year of Publication | Bias due to confounding | Bias in classification of interventions | Bias from randomisation process | Bias in. selection of participants into study | Bias due to deviations from intended interventions | Bias due to missing data | Bias arising from measurement of outcome | Risk of bias in selection of reported result | Overall |
|---|---|---|---|---|---|---|---|---|---|---|
| ROBINS-I v2 | Ngo 2024 | Serious | Low | | Low | Low | Low | Serious | Low | Serious |
| | Vitanen 2014 | Serious | Low | | Low | Serious | Low | Serious | Low | Serious |
| RoB-2 | **Winer 2023** | | | Some concerns | | Low | Low | High | Low | High |
| | **Auvinen 2022** | | | Low | | Some concerns | Low | High | Some concerns | High |
| | **Fujita 2022** | | | Low | | High | Low | High | Low | High |
| | **Winer 2022** | | | Low | | Some concerns | Low | High | Low | High |
| | **Gunvor Aasbø 2022** | | | Low | | Low | Low | High | Low | High |
| | **Sultana 2021** | | | Low | | Low | Low | High | Low | High |

### Supplemental Table 11: Quality of Included Studies Acceptability of HPV Self-sampling Screening

| Authors | Years of Publications | Country | Is the sampling frame largely representative? | Were appropriate participant recruitment methods utilized? | Is the exclusion rate acceptable? | Is the final sample size sufficient? | Are demographic variables reported? | Do the measures have adequate reliability? | Was the study conducted in a controlled setting? | Was management of data acceptable? | Overall score |
|---|---|---|---|---|---|---|---|---|---|---|---|
| Dannecker | 2004 | Germany | No | No | No | Yes | Yes | No | No | No | 2 |
| Kahn | 2005 | USA | No | No | No | Yes | Yes | Yes | No | No | 3 |
| Anhang | 2005 | USA | No | No | No | Yes | Yes | No | No | No | 2 |
| Waller | 2006 | UK | Yes | No | No | Yes | Yes | No | No | No | 2 |
| Wikstrom | 2007 | Sweeden | Yes | No | No | Yes | Yes | No | No | No | 3 |
| Barbee | 2010 | USA | No | No | No | Yes | Yes | No | No | No | 2 |
| Delere | 2011 | Germany | No | No | No | Yes | Yes | No | No | No | 2 |
| Cerigo | 2011 | Canada | No | No | No | Yes | Yes | No | No | No | 2 |
| Igidbashian | 2011 | Italy | No | No | No | Yes | Yes | No | No | No | 2 |
| Rossi | 2011 | Italy | Yes | Yes | No | Yes | Yes | No | No | No | 4 |
| Ortiz | 2012 | Puerto Rico | No | No | No | Yes | Yes | No | No | No | 2 |
| Van Baars | 2012 | The Netherlands | No | No | No | Yes | Yes | Yes | No | No | 2 |
| Bosgraaf | 2014 | The Netherlands | Yes | No | No | Yes | Yes | No | No | No | 3 |
| Galbraith | 2014 | USA | No | No | No | Yes | Yes | No | No | No | 2 |
| Virtanen | 2014 | Finland | Yes | No | No | Yes | Yes | No | No | No | 2 |
| Vanderpool | 2014 | USA | No | No | No | Yes | Yes | No | No | No | 2 |
| Castell | 2014 | Germany | Yes | Yes | No | Yes | Yes | No | No | Yes | 5 |
| Montealegre | 2014 | USA | No | No | No | Yes | Yes | No | No | No | 2 |
| Nelson | 2014 | USA | No | Yes | No | Yes | Yes | No | No | No | 3 |
| Catarino,Jr | 2014 | Switzerland | No | No | No | Yes | Yes | No | No | No | 2 |
| Chou | 2015 | Taiwan | Yes | Yes | No | Yes | Yes | No | No | Yes | 5 |
| Sultana | 2015 | Australia | No | No | No | Yes | Yes | No | No | Yes | 3 |
| Crosby | 2015 | USA | No | No | No | Yes | Yes | Yes | No | No | 2 |
| Crosby | 2016 | USA | No | No | No | Yes | Yes | Yes | No | No | 3 |

| | | | | | | | | | | | |
|---|---|---|---|---|---|---|---|---|---|---|---|
| Racey | 2016 | Canada | No | No | No | Yes | Yes | Yes | No | Yes | 4 |
| Ilangovan | 2016 | USA | No | No | No | Yes | Yes | No | No | No | 2 |
| Karjalainen | 2016 | Finland | Yes | Yes | No | Yes | Yes | No | No | No | 4 |
| Crosby | 2016 | USA | No | No | No | Yes | Yes | No | No | No | 2 |
| Levinson | 2016 | USA | No | No | No | Yes | Yes | No | No | No | 2 |
| Anderson | 2017 | USA | No | No | No | Yes | Yes | No | No | No | 2 |
| Des Marais | 2018 | USA | No | No | No | Yes | Yes | No | No | Yes | 3 |
| Molokwu | 2018 | USA | No | Yes | No | Yes | Yes | No | No | Yes | 4 |
| Kilfoyle | 2018 | USA | No | No | No | Yes | Yes | No | No | Yes | 3 |
| Adcock | 2019 | New Zealand | No | No | No | Yes | Yes | No | No | No | 2 |
| Brewer | 2019 | New Zealand | No | No | No | Yes | Yes | No | No | No | 2 |
| Reiter | 2019 | USA | No | Yes | No | Yes | Yes | No | No | No | 3 |
| Malone | 2020 | USA | Yes | Yes | No | Yes | Yes | Yes | No | Yes | 6 |
| Datta | 2020 | Canada | No | No | No | Yes | Yes | No | No | Yes | 3 |
| Andersson | 2021 | Sweden | No | No | No | Yes | Yes | No | No | No | 3 |
| Bromhead | 2021 | New Zealand | No | No | No | Yes | Yes | Ni | No | Yes | 4 |
| Veerus | 2021 | Estonia | Yes | Yes | No | Yes | Yes | No | No | No | 4 |
| Smith | 2022 | USA | No | No | No | Yes | Yes | No | No | Yes | 3 |
| Chaw | 2022 | Brunei | No | No | No | Yes | Yes | No | No | No | 2 |
| Ngu | 2022 | Hong Kong | No | No | No | Yes | Yes | No | No | No | 2 |
| Sherman | 2022 | New Zealand | Yes | No | No | Yes | Yes | No | No | No | 3 |
| Parker | 2022 | USA | No | Yes | No | Yes | Yes | No | No | No | 3 |
| Zhu | 2022 | Canada | No | No | No | Yes | Yes | No | No | Yes | 3 |
| Fujita | 2023 | Japan | No | No | No | Yes | Yes | No | No | Yes | 3 |

Negative agreement was also affected by setting of the test (p <0.001) and the test positivity rate ratio was jointly affected by self-sampling device and assay method (p <0.001) (*Table 4)*. Other outcomes were not affected by the other characteristics tested.

### Supplemental Figure 1: Overall Agreement

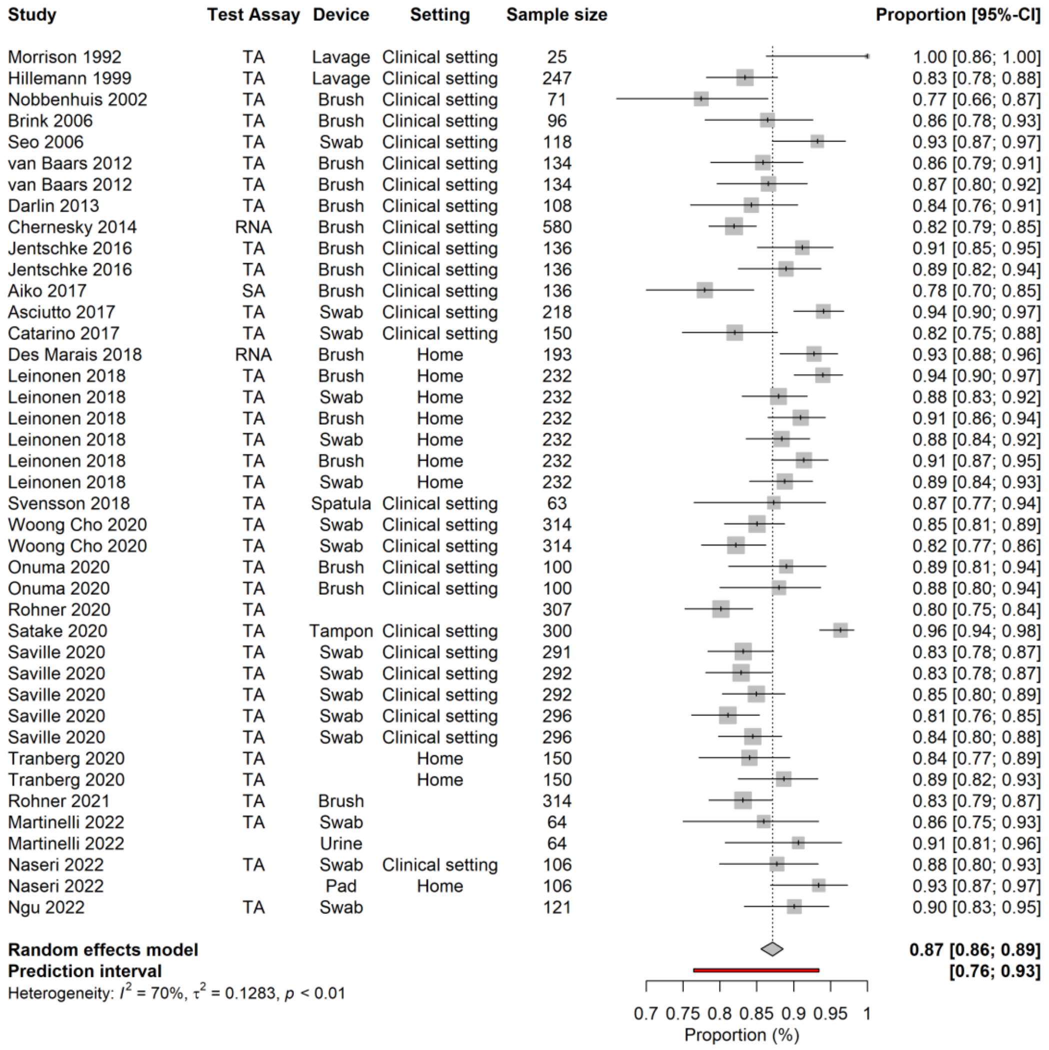

### Supplemental Figure 2: Forest plot for kappa

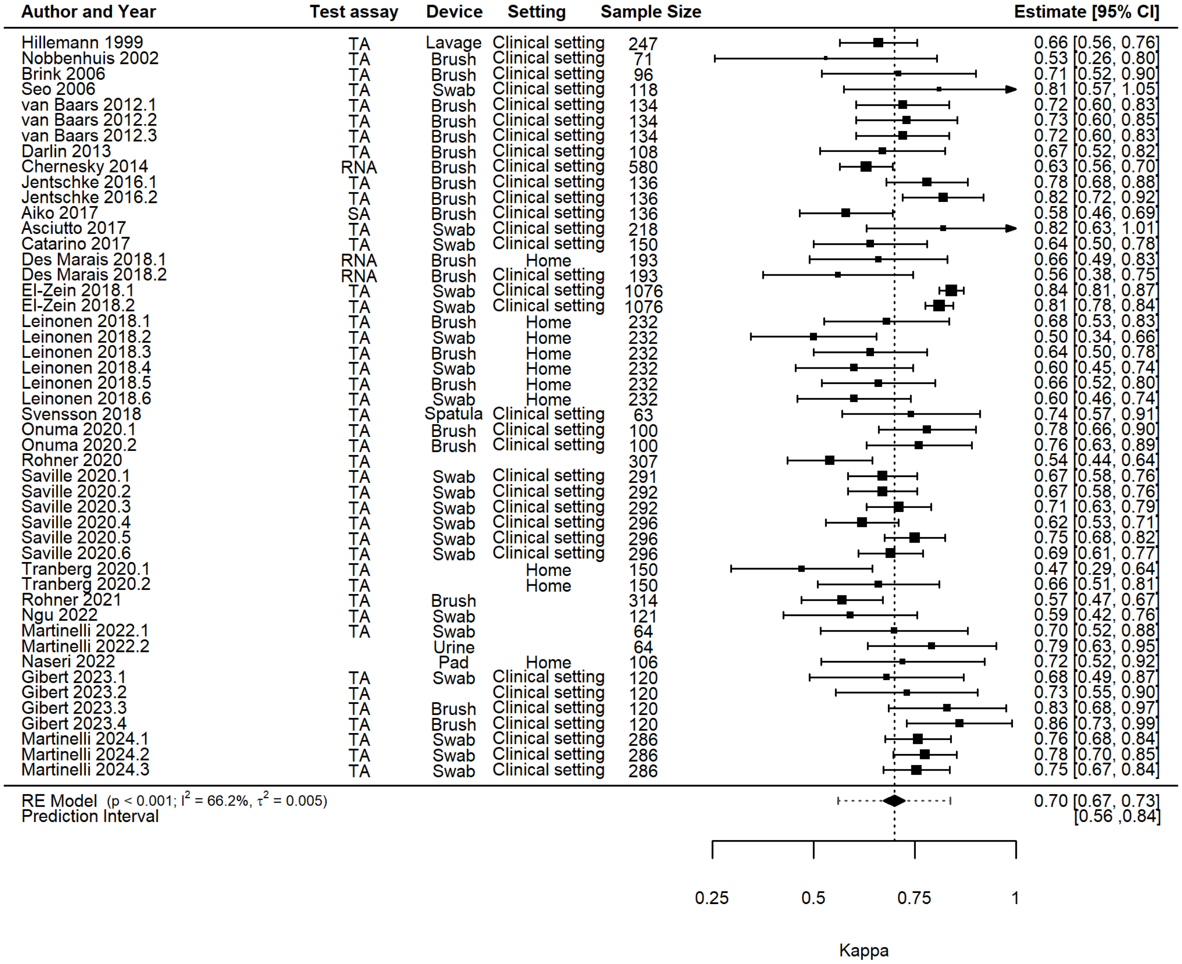

### Supplemental Figure 3: Overall Agreement across Settings

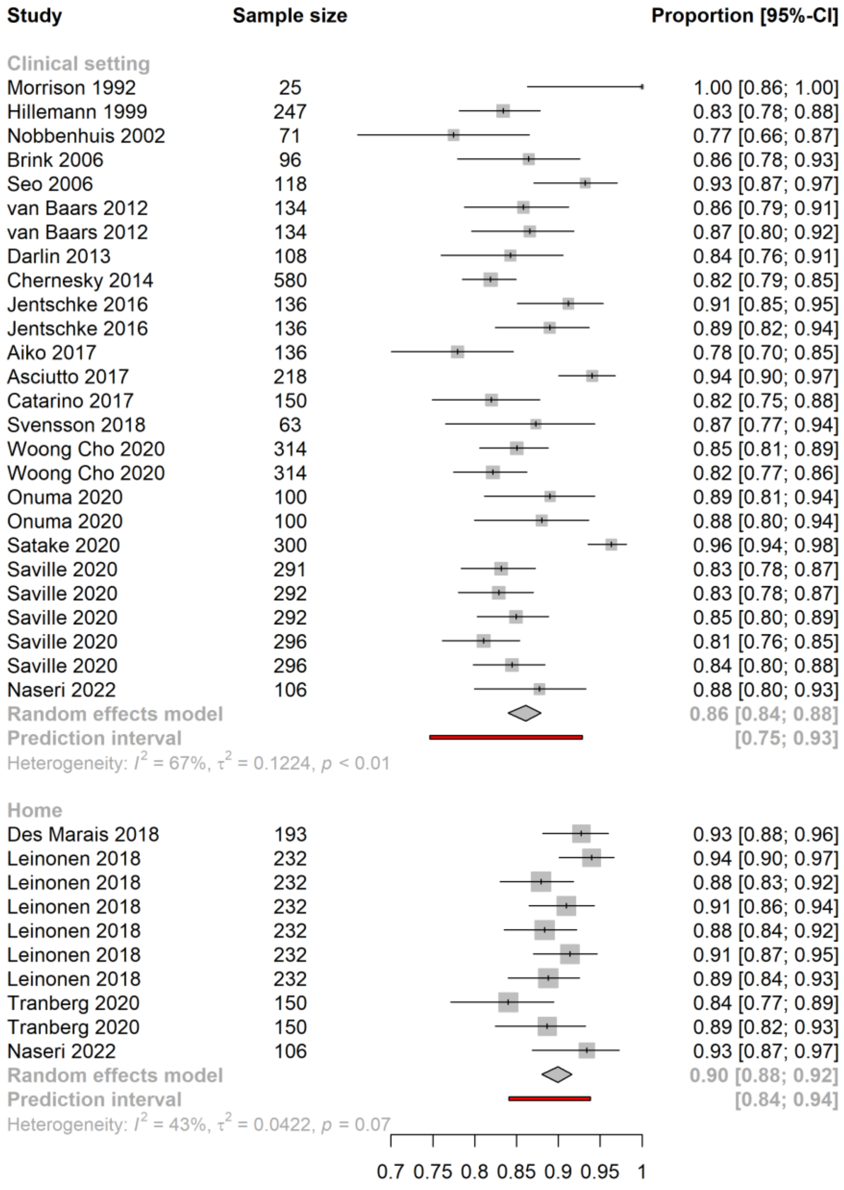

### Supplemental Figure 4: Kappa by setting

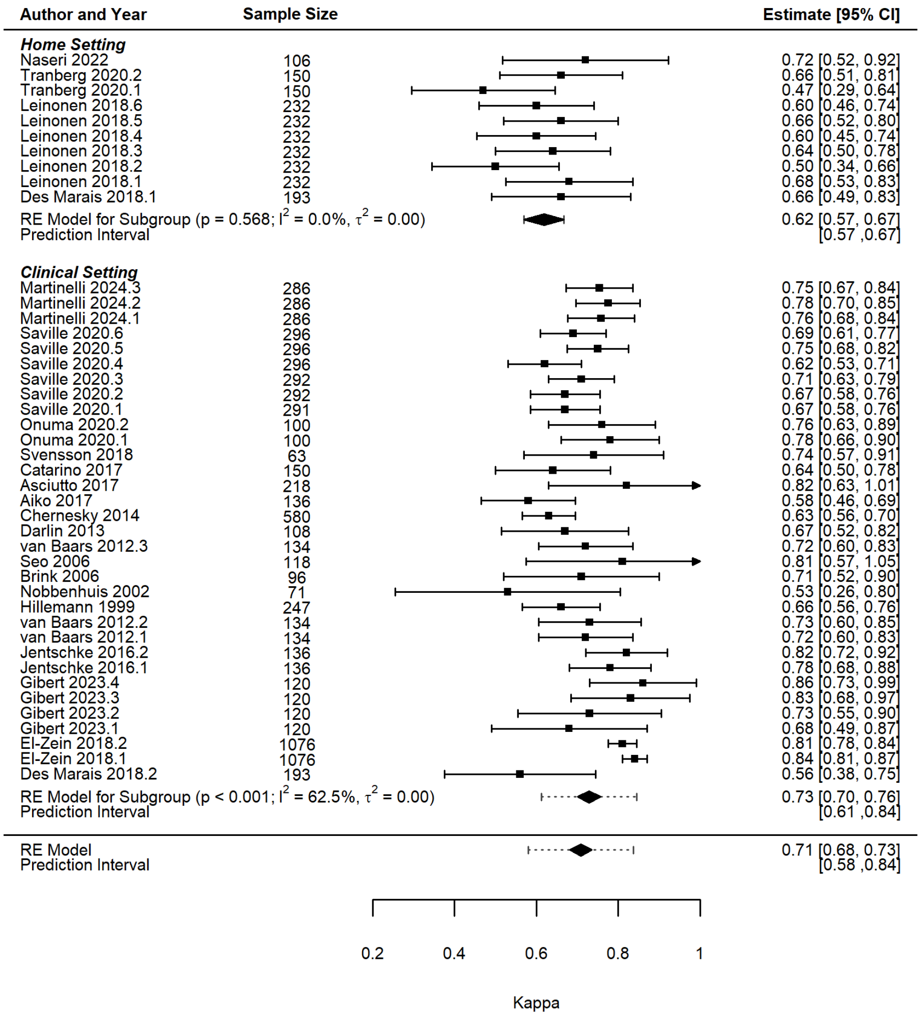

### Supplemental Figure 5: Difference in Participation Rate between Self-sampling and Control

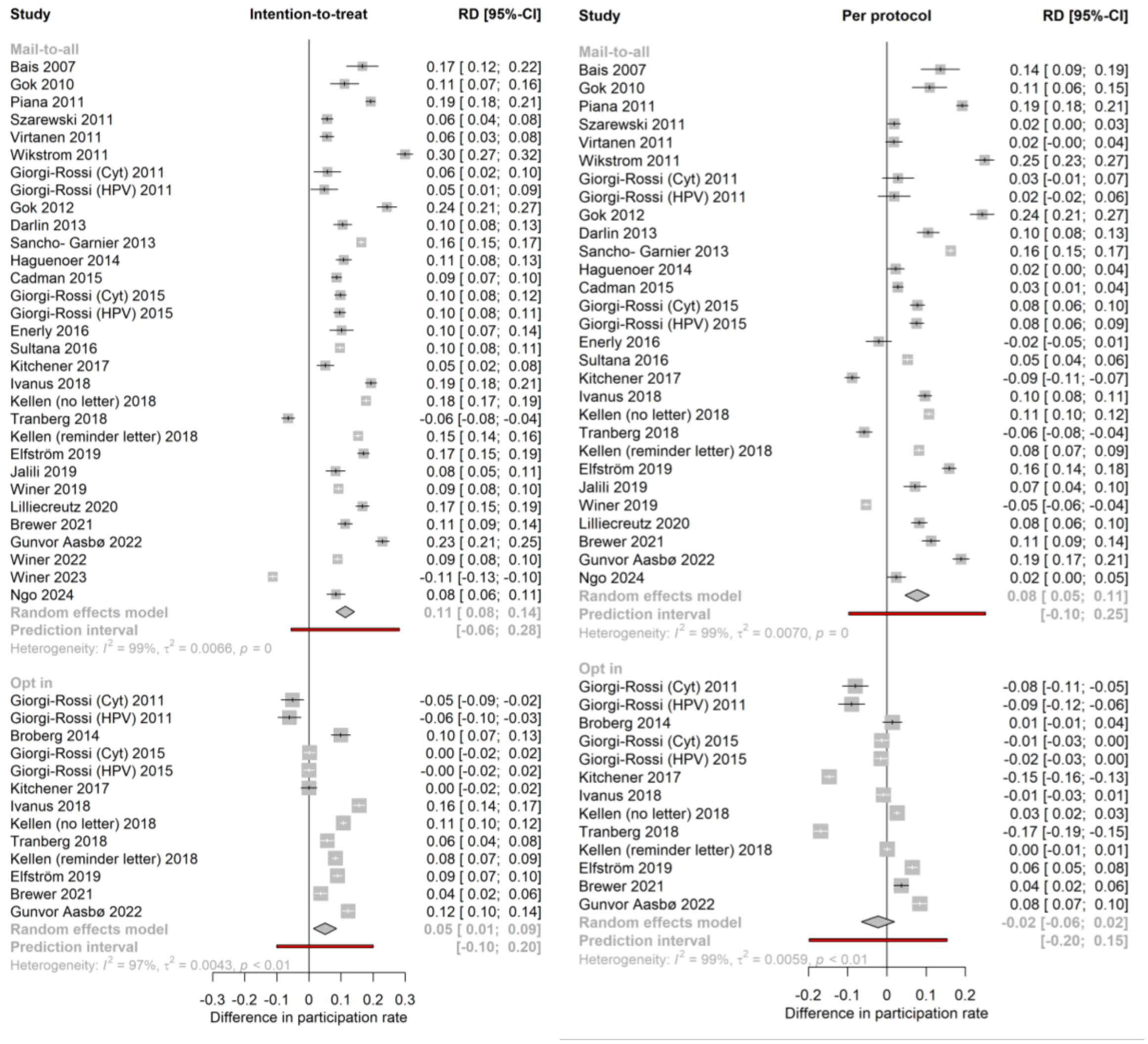

### Supplemental Table 12: Sample adequacy, adherence, and CIN2+ detection rates

| Parameter | No. of studies | Absolute proportion self-sampling (% unless other specified) (95% CI) |
|---|---|---|
| Unsatisfactory sample | 20 | 0.9 (0.6 to 1.2) |
| Adherence to follow-up | 29 | 80.5 (72.2 to 86.7) |
| CIN2+ detection (per thousand women screened) | 25 | 11.6 (8.4 to 16.0) |

### Supplemental Figure 6: General Acceptability of Self-sampling

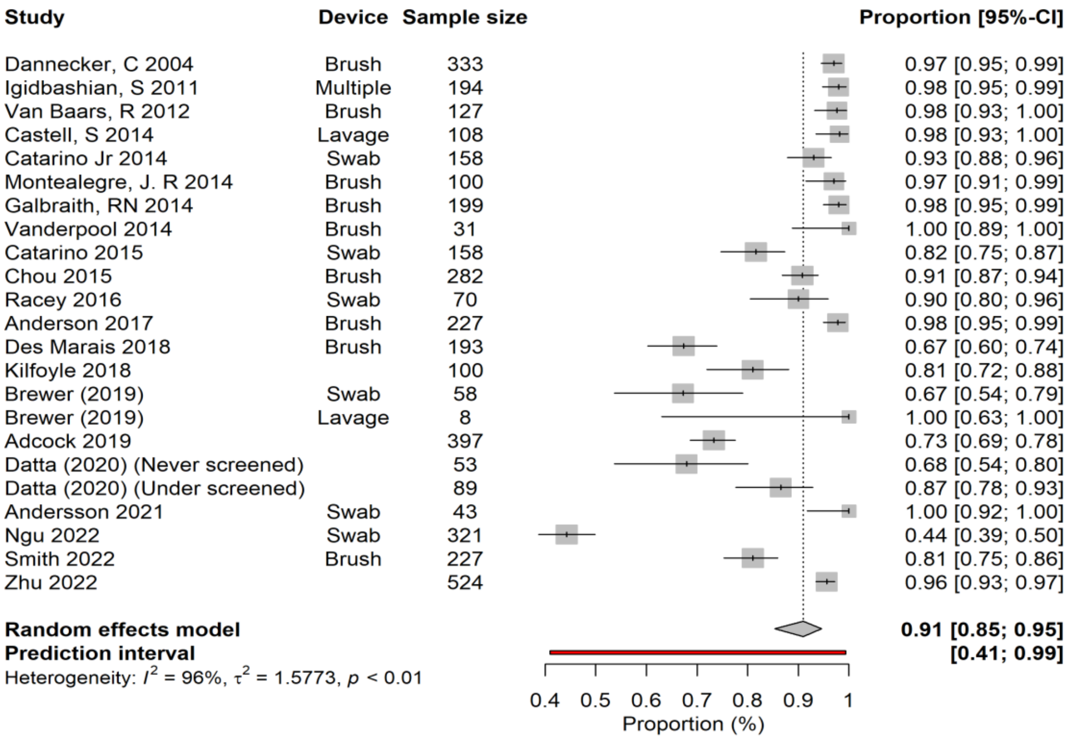

### Supplemental Figure 7: Women Preferring Self-sampling to Healthcare Professional Sampling

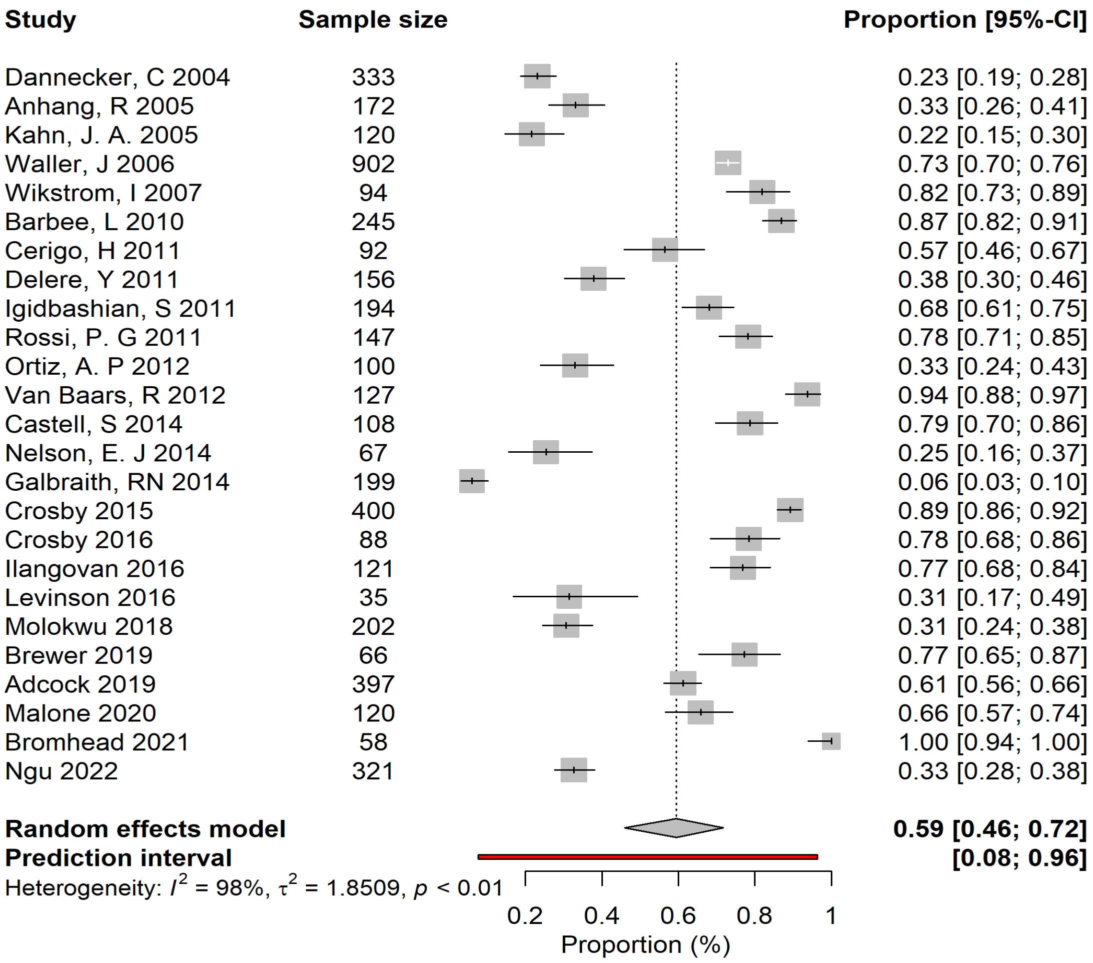

**Supplemental Figure 8: Stated Preference for Self-sampling at Home versus Healthcare Setting According to Sample Device**

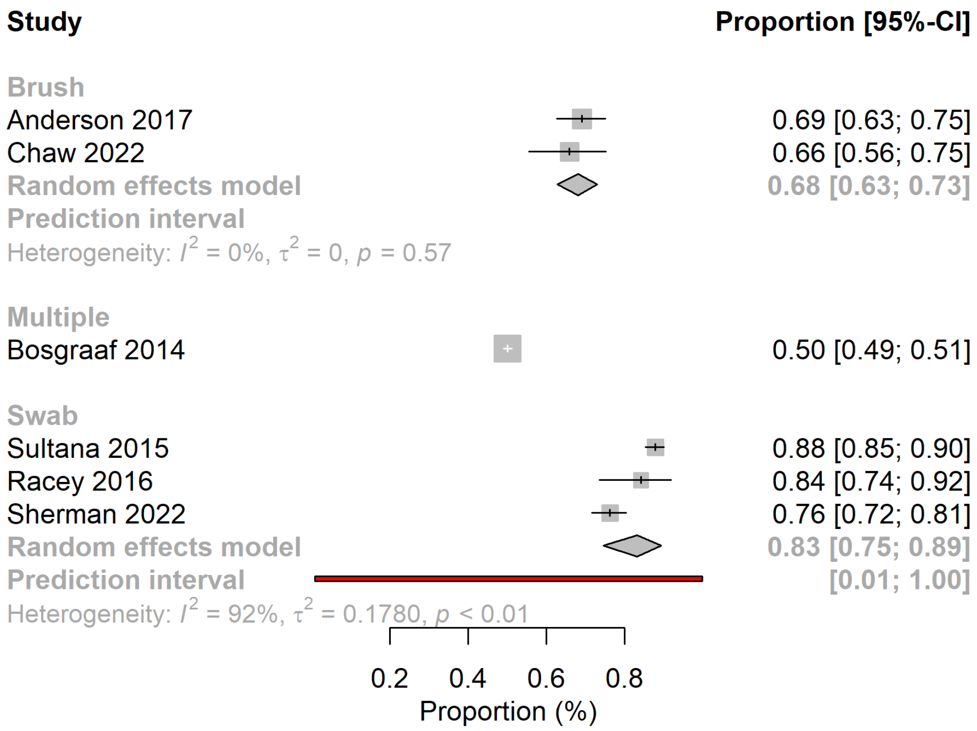

**Supplemental Figure 9: Stated Preference for Self-sampling at Home versus Healthcare Setting According to Invitation Strategy**

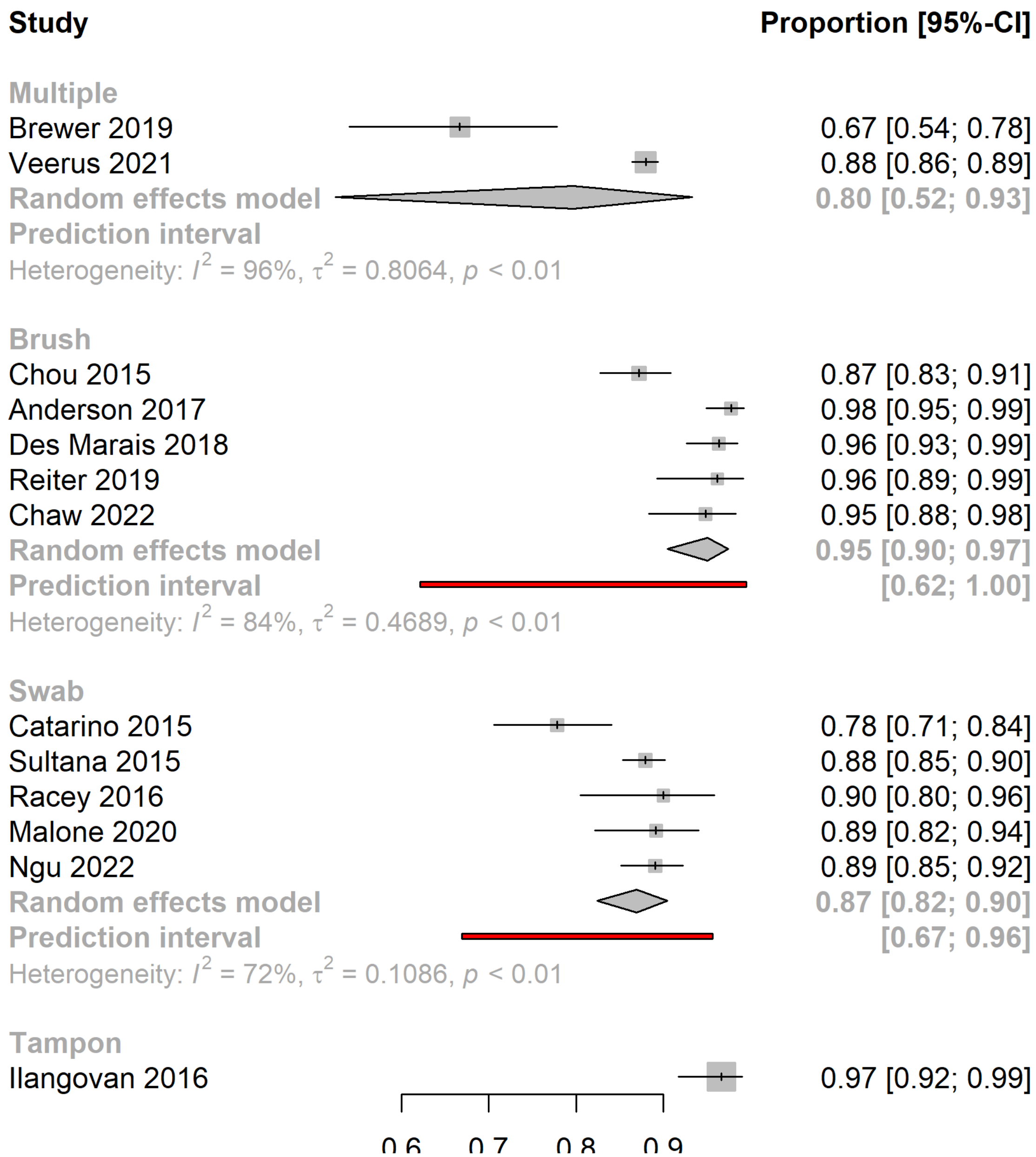

### Supplemental Figure 10: Stated Willingness to Repeat Cervical Screening

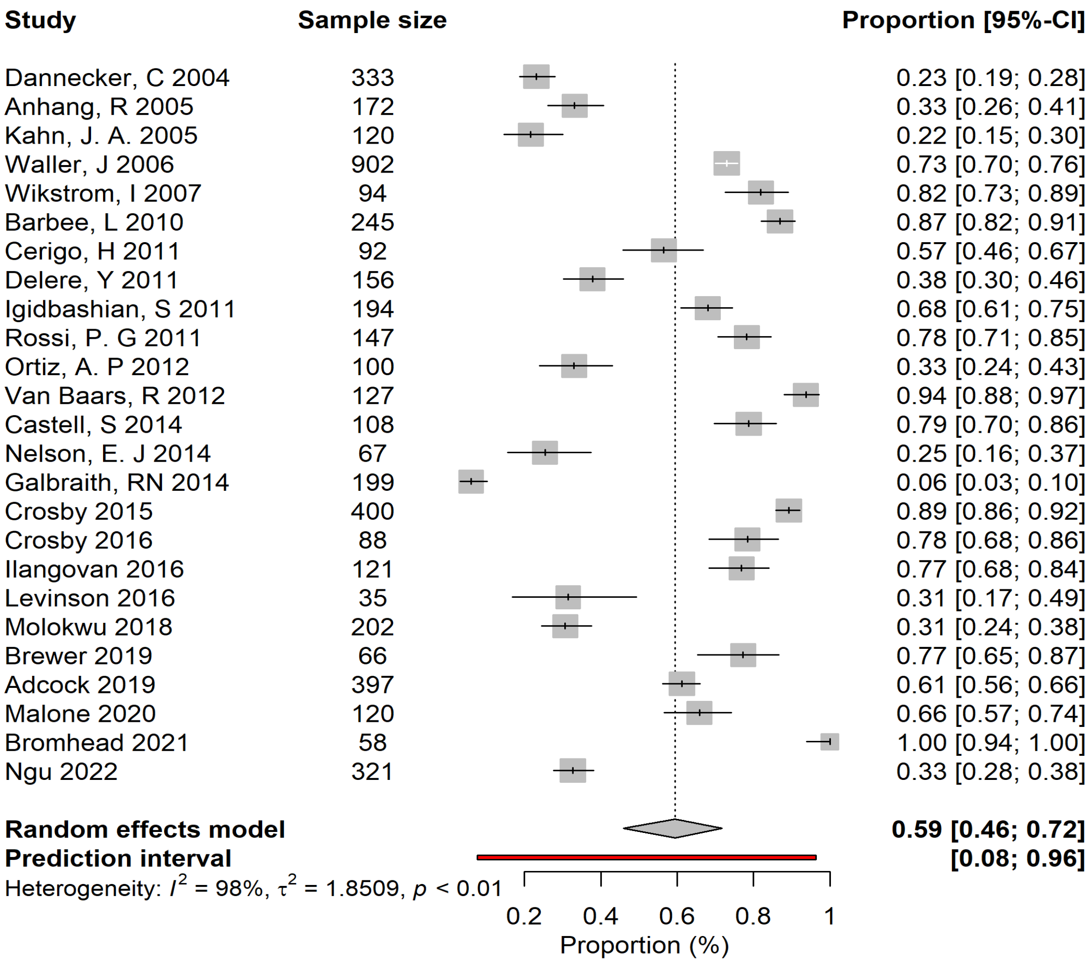

48. Ardashel Latsuzbaia, Severien Van Keer, Davy Vanden Broeck, Steven Weyers, Gilbert Donders, Philippe De Sutter, Wiebren Tjalma, Jean Doyen, Alex Vorsters and MA. Clinical Accuracy of Alinity m HR HPV Assay on Self- versus Clinician-Taken Samples

Using the VALHUDES Protocol. 2023;

75. Gok M, Heideman DAM, Folkert J van Kemenade, Johannes Berkhof LR, WM J, Spruyt, Feja Voorhorst, Jeroen AMBelie¨n, Milena Babovic 4 Peter J F Snijders, Meijer CJLM. HPV testing on self collected cervicovaginal lavage specimens as screening method for women who do not attend cervical screening: cohort study. :1–8.

96. Winer RL, Lin J, Tiro JA, Miglioretti DL, Beatty T, Gao H, et al. Effect of Mailed Human

Papillomavirus Test Kits vs Usual Care Reminders on Cervical Cancer Screening Uptake , Precancer Detection , and Treatment A Randomized Clinical Trial. 2019;2(11):1–14.

107. Winer RL, Lin J, Anderson ML, Tiro JA, Green BB, Gao H, et al. Strategies to Increase

Cervical Cancer Screening With Mailed Human Papillomavirus Self-Sampling Kits A Randomized Clinical Trial. 2024;98195.

108. Ito Taro TO, , Tetsuji Kurokawa , Yoko Chino AS and YY. Evaluating Opt-In Vaginal Human Papillomavirus Self-Sampling : Participation Rates and Detection of High-Grade. 2024;

115. Kahn JA, Slap GB, Huang B, Rosenthal SL, Wanchick AM, Kollar LM, et al. Comparison of Adolescent and Young Adult Self-Collected and Clinician-Collected Samples for Human Papillomavirus.

116. R. Anhang, J.A. Nelson, R. Telerant, Chiasson, And Thomas C. Wright J. Acceptability of Self-Collection of Specimens for HPV DNA Testing in an Urban Population. J Women's Heal. 2005;14(8):721–8.

117. Waller J, Mccaffery K, Forrest S, Szarewski A, Cadman L, Austin J, et al. Acceptability of unsupervised HPV self-sampling using written instructions. 2006;13(4):208–13.

118. Ingrid Wikstrom HS& EW. Attitudes to self-sampling of vaginal smear for human papilloma virus analysis among women not attending organized cytological screening. Acta Obstet Gynecol. 2007;(February):720–5.

119. Barbee L, Kobetz E, Menard J, Cook N, Blanco J, Barton B, et al. Assessing the acceptability of self-sampling for HPV among Haitian immigrant women : CBPR in

action. 2010;421–31.

120. Cerigo H. HPV Knowledge and Self-Sampling for the Detection of HPV DNA among Inuit women in Nunavik , Quebec. 2010;(August).

142. Kilfoyle KA, Marais AC Des, Ngo MA, Romocki L, Richman AR, Barclay L, et al. Preference for Human Papillomavirus Self- Collection and Papanicolaou: Survey Of

Underscreened Women in North Carolina. J Low Genit Tract Dis. 2019;22(4):302–10.

143. Jennifer C. Molokwu, Eribeth Penaranda, Alok Dwivedi, Indika Mallawaarachchi and N. 22571302 QCL :: University of Glasgow QCL :: University of Glasgow 227874. 2024;

154. Sherman SM, Psychology R, Brewer N, Mbchb KB, Outcomes H, Bromhead C, et al. Human papillomavirus self - testing among unscreened and under - screened M ā ori ,

Pasifika and Asian women in Aotearoa New Zealand : A preference survey among responders and interviews with clinical - trial nonresponders. 2022;(August):2914–23.

155. Zhu P, Tatar O, Haward B, Gri G, Perez S, Smith L, et al. Assessing Canadian women ’ s preferences for cervical cancer screening : A brief report.

156. Id MF, Id KN, Shimazu M, Suzuki M. Acceptability of self-sampling human papillomavirus test for cervical cancer screening in Japan : A questionnaire survey in the ACCESS trial. 2023;196:1–15.
